# Profiling Neoadjuvant Therapy Response in Rectal Cancer Using Publicly Available Transcriptomic RNA-seq Datasets

**DOI:** 10.1101/2025.07.18.25331799

**Authors:** Aleksandra Stanojevic, Rafael Stroggilos, Mladen Marinkovic, Ana Djuric, Suzana Stojanovic-Rundic, Radmila Jankovic, Sergi Castellvi-Bel, Remond J.A. Fijneman, Antonia Vlahou, Jerome Zoidakis, Milena Cavic

## Abstract

Neoadjuvant chemoradiotherapy followed by total mesorectal excision is standard for locally advanced rectal cancer, but response varies and current markers are insufficient. This study integrates public bulk RNAseq data to identify predictive features of response. *TRIM54* and *PABPC4* were up-regulated in the responder group, while *ADSS1* and *MGAT1* were up-regulated in non-responder group. *ARMC2* was identified as a predictive biomarker up-regulated in pathological complete response. Responder group showed enrichment of NK cells and CD4+ lymphocytes, while immune precursors were linked to poor outcome. Transcription factor analysis revealed *SP1* and *NFKB* activations in the non-responder group and *TCF15* in responder group. *SMAD3* and *RDXANK* were associated with complete regression, while *MYC* was dominant in incomplete regression. These findings provided insight into mechanisms underlying therapy response. To our knowledge, this is the first meta-analysis using high-throughput sequencing data, providing a valuable starting point for future rectal cancer research.

## INTRODUCTION

Colorectal cancer is the third most prevalent form of cancer globally in both sexes with nearly two million newly diagnosed cases in 2022, according to Globocan statistics ^1^. Although colorectal cancer has been considered as a one disease entity for a long time, it is now acknowledged that colon cancer and rectal cancer represent two distinct types of disease at pathological, clinical and molecular levels ^2–4^. Increasing evidence suggests that these two cancer types should be further divided based on their molecular profile on both genomic and transcriptomic levels thus classifying them into different molecular subtypes while considering their inherent heterogeneity ^3–6^. Rectal cancer (RC) represents approximately one-third of total colorectal cancer cases ^7,8^ while locally advanced rectal cancer (LARC) is characterized as stage II (T3/4N0M0) and III (T1-4 N+ M0) ^9^.

Since 2004, neoadjuvant chemoradiotherapy (nCRT) has been the standard of care for locally advanced rectal cancer (LARC). This treatment typically consists of chemotherapy based on 5-fluorouracil combined with radiation therapy at a dose of 50.4–54 Gy, followed by total mesorectal excision (TME), with or without adjuvant chemotherapy ^8^. Approximately 10–20% of patients achieve a pathological complete response (pCR) after TME, defined as the absence of residual tumor cells in the surgical specimen (ypT0N0) ^10–12^. However, around 35% of patients show a poor or no response to nCRT, and a smaller subset, estimated at 2–4%, may even experience distant progression during treatment^13–15^.

In some cases, patients achieve a clinical complete response (cCR) after nCRT. This is defined as the absence of residual tumor upon clinical reevaluation, which includes digital rectal examination, pelvic magnetic resonance imaging (MRI), and proctoscopy. For patients with cCR, a non-operative management strategy known as the “watch-and-wait” approach may be considered, involving close follow-up and reserving surgery only in cases of local tumor regrowth. However, the concordance between cCR and pCR remains low ^16,17^.

To improve cCR rates following neoadjuvant treatment, current research directions focus on several key strategies: prolongation of the interval between the completion of neoadjuvant therapy and surgery, escalation of radiotherapy dose, and intensification of systemic treatment within the framework of total neoadjuvant therapy (TNT) ^18–22^. Furthermore, the incorporation of immunotherapy in the management of mismatch repair-deficient (dMMR) rectal cancer has expanded the therapeutic landscape and opened new possibilities for personalized treatment ^23^. Within the organ preservation paradigm, particularly in patients demonstrating a clinical complete response, the watch-and-wait approach aims to maintain oncologic safety while preserving quality of life. These evolving therapeutic modalities underscore the increasing need for refined patient stratification and risk-adapted treatment strategies to optimize outcomes.

Despite significant variability in patient responses, neoadjuvant chemotherapy remains a conventional treatment protocol. At the molecular level, currently, there is still a lack of clinically validated predictive features that can distinguish treatment response and enable improved quality of life. Identifying predictive biomarkers will enable improved patient selection and enhance treatment optimization opportunities. Furthermore, the watch-and-wait approach still has limited potential, as the primary objective of organ preservation may be compromised by the potential for tumor regrowth, distant metastasis and ultimate local failure ^24^. Discovery of predictive biomarkers may improve patient selection with clinical complete responses that could be eligible for a watch-and-wait approach and prevent TME or, at the very least, prolong the interval between treatment and surgery.

It is widely recognized that the cellular response to DNA damage induced by irradiation is complex and involves multiple forms of cell death including apoptosis, mitotic devastation, senescence, necrosis, autophagy and stress signaling pathways, whereas chemotherapy serves to sensitize tumor cells to irradiation. Furthermore, it has been reported how radiation affects tumor cells and the surrounding during treatment but there is a lack of evidence regarding the impact of transcriptomic and proteomic profiles of tumor and tumor-microenvironment (TME) on tumor’s therapeutic response ^25–27^.

Transcriptomic profiling of tissue provides characterization of molecular features and enables insight into characteristics that can be used for better understanding of mechanism behind response. LARC was profiled through various omics approaches and technologies ^28–32^. Research conducted by our team demonstrated that patients with varying responses exhibit distinct proteomics profiles ^33^ as well as hematological features ^34^ that can be used for their stratification. By applying a radiomics approach, radiomic features showed differences between treatment outcomes ^35^. Numerous studies examining LARC tissue have been performed at various levels but most suffer from the limitation of small sample size, resulting in limited statistical power and usually the lack of validation of results obtained. Conversely, the integration of studies conducted at the same omics level may enhance statistical power and increase likelihood of significant biological findings.

This study aims to integrate publicly available bulk RNAseq data and as a step beyond the state of the art in this disease context, to perform a comprehensive characterization of LARC at the trascriptome level, towards the identification of predictive biomarkers. To date, meta-analyses concerning LARC were conducted using microarray transcriptomic data^36^; to the best of our knowledge this represents the first study that uses publicly available transcriptomic data derived from Next Generation Sequencing methodologies, offering comprehensive insights.

## MATERIALS AND METHODS

### Study cohort

A comprehensive dataset search strategy was employed, encompassing the Gene Expression Omnibus and PubMed repositories using a variety of keywords as shown in Figure 1 conducted on 31/07/2024. Diverse range of keywords were used to include all articles/datasets containing transcriptome RNAseq data on pretreatment biopsy specimens along with data on therapeutic response to therapy after surgery of LARC patients. Studies examining other anatomical segments of the colorectum, as well as those conducted on animal models or cell lines were excluded. The inclusion and exclusion criteria are presented in Figure 1. Throughout the search process, overlapping results were noted among different keywords. Following extensive searches for rectal cancer studies, across Gene Expression Omnibus (GEO) and PubMed repositories five transcriptomics datasets were identified fulfilling the inclusion criteria, encompassing a total of 248 samples (GSE190826^37^, GSE233517^38,39^, GSE209746 ^40^, GSE80606 ^41^, Toomey et al.^42^) as detailed in Table 2-3, Figure 1. All patients were diagnosed with LARC and underwent nCRT utilizing 5-FU or Capecitabine, some patients were treated in accordance with the total neoadjuvant therapy (TNT) model which incorporates oxaliplatin, either induction or consolidation. Patient responses were evaluated after surgery using different methods as shown in Table 1. The scales were standardized ^43^ and the patients were categorized into responders (R) and non-responders (NR) or alternatively as those exhibiting a pathological complete response (pCR) and pathological non-complete response (Non pCR) Table 1. Patients classified as responders (R) are defined as those achieving either complete response or a near-complete response to therapy. Patients exhibiting absent or minimal response were classified as nonresponders (NR). The same patient cohort was stratified into those achieving a pathological complete response (pCR), characterized by non-detectable tumor cells after surgery, and all others achieved pathological non-complete response (Non pCR).

**Figure 1.**
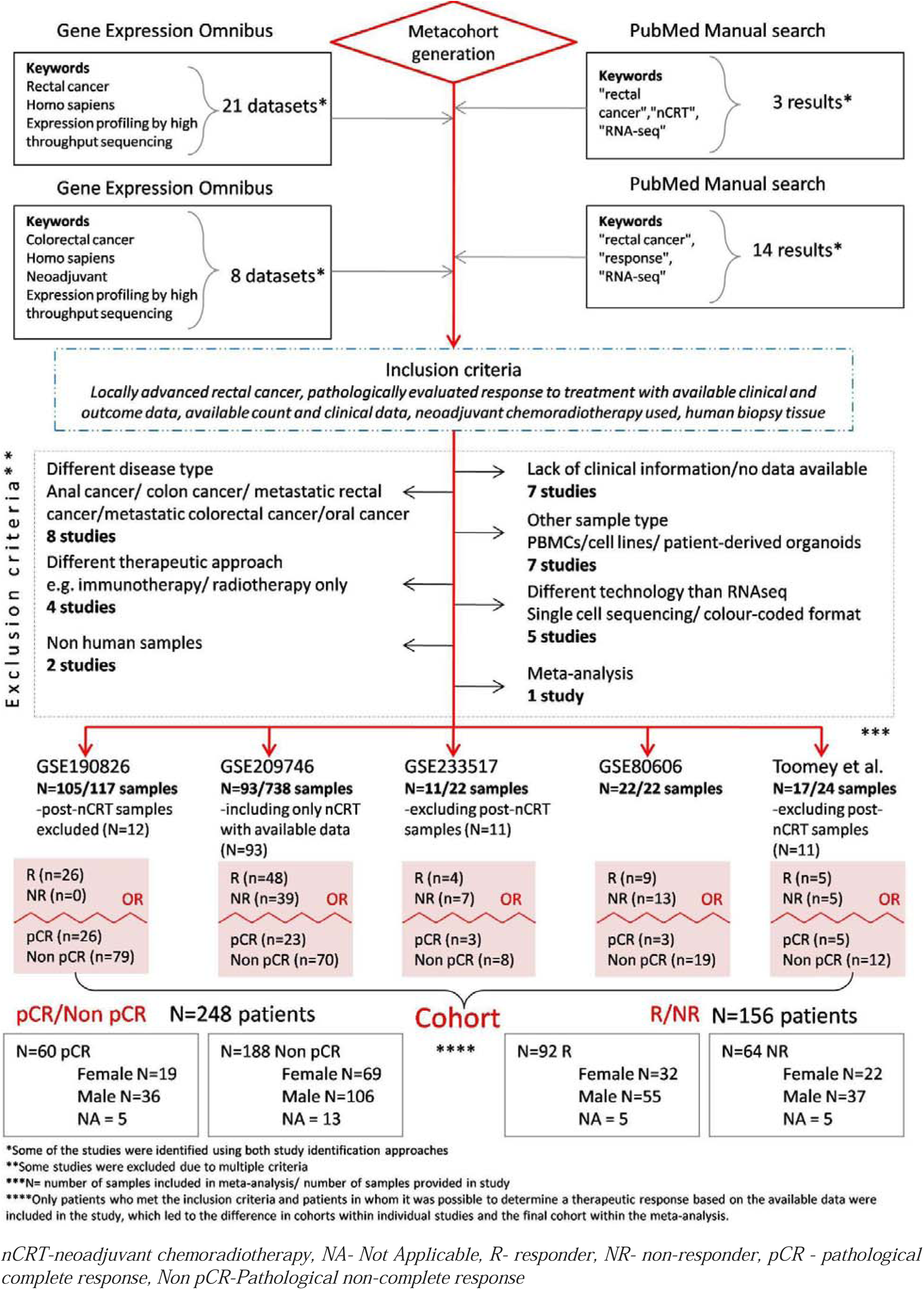
Transcriptomic data search flowchart. The publicly available scientific database PubMed together with the Gene Expression Omnibus database was used to search for studies conducted on pretreatment LARC samples with the aim of determining predictive biomarkers. Studies were reviewed based on the inclusion criteria. The final group consisted of five studies where patients were divided into responders (TRG1-2, Mandard scale) and non-responders (TRG3-5, Mandard scale) or pCR (TRG1, Mandard scale) and Non pCR (TRG2-5, Mandard scale)

**Table 1.** Harmonization of different classifications of response to nCRT.

| Mandard five-tiered TRG | Dworak five-tiered system | CAP System | RCPath System | Ryan modified three-tiered Mandard TRG | Responder (R)/Nonresponder (NR) | Pathological complete responders (pCR)/ Non pathological complete responders (Non pCR) |
| --- | --- | --- | --- | --- | --- | --- |
| TRG 1 | TRG 4 | TRG 0 | RCPath A | TRG 1 | R | pCR |
| TRG 2 | TRG 3 | TRG 1 | RCPath B |  | R | Non pCR |
| TRG 3 | TRG 2 | TRG 2 | RCPath C | TRG 2 | NR | Non pCR |
| TRG 4 | TRG 1 | TRG 3 |  | TRG 3 | NR | Non pCR |
| TRG 5 | TRG 0 |  |  |  | NR | Non pCR |

*nCRT- neoadjuvant chemoradiotherapy, TRG- tumor regression grade, R- responder, NR- non-responder, pCR - pathological complete response, Non pCR- Pathological non-complete response, RCPath- Royal College of Pathologists, CAP- College of American Pathologists*

**Table 2.**
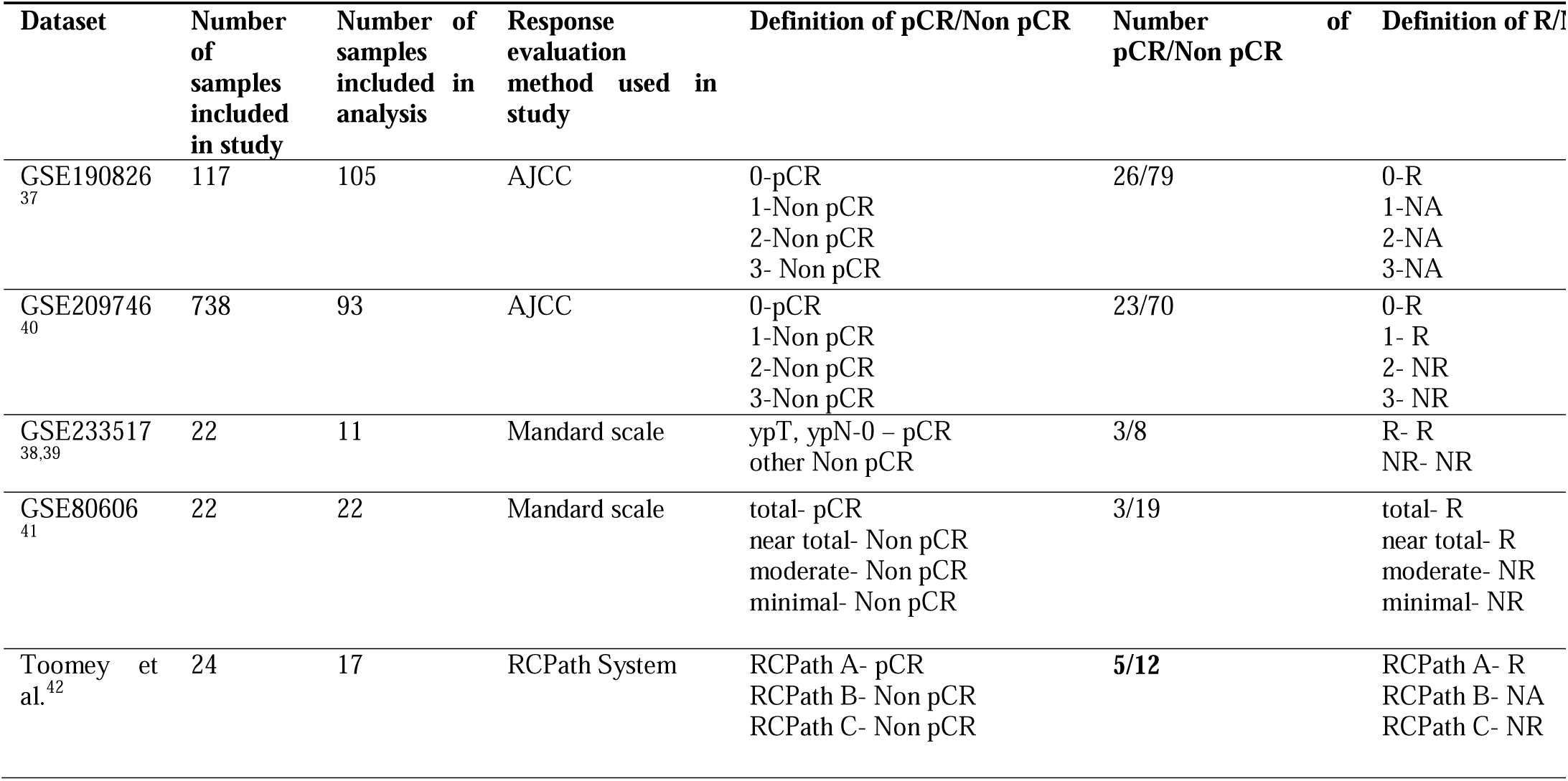
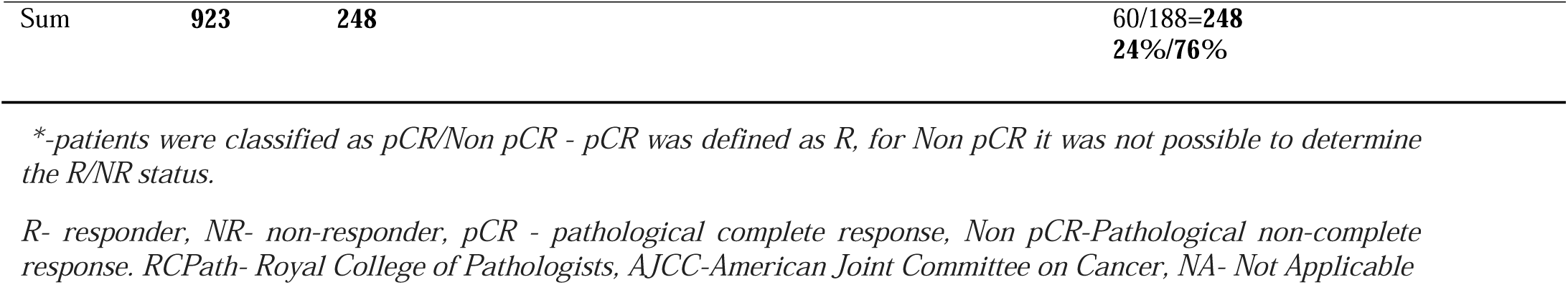
Transcriptomics datasets characteristics.

### Combined batch-free dataset generation and Differential gene expression analysis

Each dataset was initially analyzed independently to identify differentially expressed genes (DEGs) between R/NR and pCR/NonpCR. Genes exhibiting a cumulative count of less than 10 across samples were excluded. Differential expression analysis was performed using the DESeq2 v1.42.1 pipeline ^45^, specifically developed for the analysis of RNAseq raw counts data. Statistical analysis was performed by applying the Wald statistics, while p-values were corrected with Benjamini-Hochberg adjustment method for multiple comparisons. GSE190826 was excluded from the individual R and NR comparison due to absence of NR data. Beside the identification of predictive biomarkers for R/NR, predictive biomarkers of pathological complete response were identified. The combined cohort was generated by integrating samples from five datasets based on genes present in all studies (intersection). Genes with redundant sequencing data were retained by selecting those with the highest mean counts across all samples. Given that different generations of sequencing technology were used (Illumina NextSeq 500, Illumina HiSeq 2000,

Illumina HiSeq 2500, and Illumina HiSeq 4000), and the variation in sample and library sizes across datasets, batch effect removal was performed using the ComBat-seq algorithm (sva v3.35.2 ^46^) and DESeq2 for downstream differential expression analysis. ComBat Seq is a variation of ComBat method grounded in empirical Bayes techniques and, in contrast to alternative batch adjustment methods, assumes that data follows a negative binomial distribution. In addition, this approach ensures a consistent workflow for both individual datasets and the combined cohort while maintaining the negative binomial distribution in the batch-adjusted counts data and preserving integer counts form. ComBat seq effectively reduces batch effects by eliminating technical variances and preserving biological variability (Figure 3). A comprehensive overview of the methodology employed in this meta-analysis is provided in Figure 2. After obtaining a representative combined dataset, DESeq2 pipeline with the dataset as covariance was applied in order to identify DEGs between R/NR and pCR/Non pCR.

**Figure 2.**
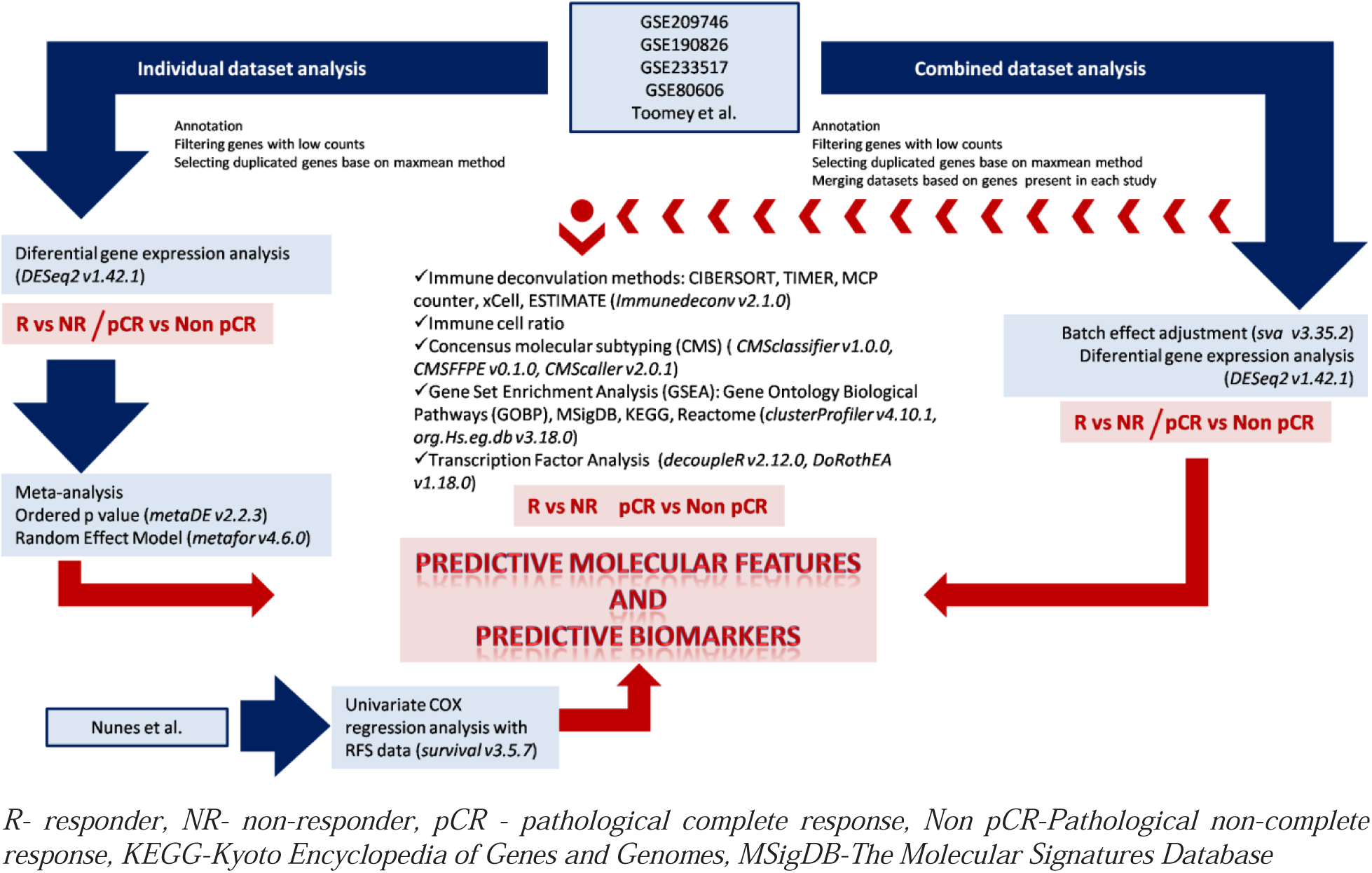
Overview of the methodological approach used to identify predictive parameters of response (R vs NR and pCR vs Non pCR) to neoadjuvant chemoradiotherapy in patients with LARC.

**Figure 3.**
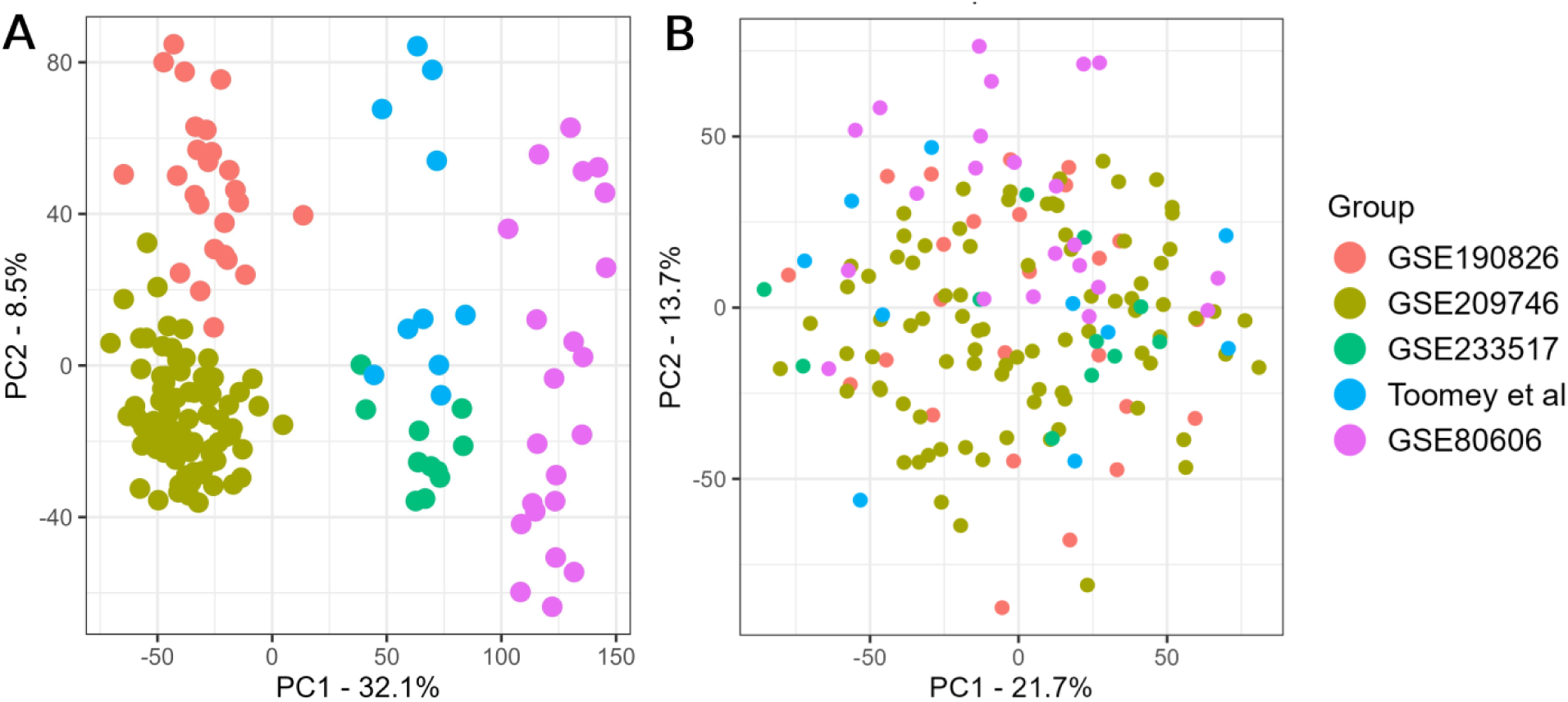
PCA plot before (A) and after (B) batch correction. PCA, Principal component analysis.

### Meta-analysis of individual datasets

Meta-analysis approaches, ordered p-value (rOP) method and the random effect model (REM) was used to identify predictive biomarkers of response to nCRT in LARC based on statistical output of individual studies. ^47^ OrP method accounts for the distribution of p value for each gene across analyzed datasets, whereas REM approach assesses the LogFC by considering the distribution of effect sizes for each gene, accounting for inter- and intra-study variations, thereby estimating the effect size along with the corresponding p-value and adjusted p-value. In order to consider both effects in meta-analysis setting methods, p-value and Log2FC from all analyzed genes in individual datasets were used. The estimation of between-study variance was performed employing the Restricted Maximum Likelihood (REML) method. To perform meta-analysis, R packages MetaDE v2.2.3 ^48^ and metafor v4.6.0^49^ were used.

### Validation cohort

The study of Nunes et al^44^ published in August 2024 was used for independent validation, in which the level of gene expression as prognostic markers of LARC was examined. No independent study examining the predictive potential of gene expression was available at the time of validation. Regardless, it is expected that patients who have a better response to therapy have a longer survival without tumor regression and vice versa. Patients with available RNAseq data who had stage II and III rectal cancer and received nCRT were subtracted from the study. A total of 68 patients (33 female, 35 male) with a median age of 67 (range 38-93) were included in the validation study. Regression-free survival (RFS) was used as an input variable and was defined as the interval from surgery to the earliest incidence of local or distal recurrence or mortality. Univariate COX regression analysis was performed to evaluate the prognostic impact of gene expression using the R package survival v3.5.7 ^50^. The results of COX analysis were used to validate the identified predictive markers, validation criteria was the direction of the hazard ratio (HR). RFS curves of predictive biomarkers were generated using the Kaplan-Meier method from the survminer v0.5.0 ^51^ package and patients were categorized based on the median gene expression into “low” and “high”.

### Biomarker discovery

To identify predictive biomarkers associated with response to neoadjuvant Chemoradiotherapy (nCRT), outputs from differential expression analysis of both the combined dataset and meta-analysis (orP and REM) settings were systematically compared. A gene was identified as a predictive marker if it fulfilled the following criteria: differentially expressed genes (DEGs) with p-value <0.05 and adjusted *p*-value <0.25 in the combined dataset, REM p-value <0.05, and orP p-value< 0.05. The predictive efficacy of identified biomarkers was confirmed or denied through the analysis of RFS data as an independent validation, employing COX regression analysis with p value<0.05 and HR with the same trend as z value of REM and LFC in DESeq2.

### Immune cells deconvolution

Immune deconvolution analysis employing MCPcounter, ESTIMATE, xCelll, CIBERSORT and TIMER method from the immunedeconv package v2.1.0 ^52^ by using TPMs data prior batch effect correction were conducted. MCPcounter and TIMER facilitated the quantification of immune infiltration within tumor tissue, ESTIMATE was used for the assessment of tumor purity, as well as the immune and stromal score. xCell was utilized for the relative infiltration of 64 distinct types of immune and stromal cells, alongside CIBERSORT. Furthermore, in addition to the overrepresentation of specific immune subtypes statistical significance of the immune cells ratio across different treatment outcomes was investigated. During the ratio computation, all ratios that had the value NaN, Inf and 0 were classified as NAs. Ratios exceeding 80% NAs were excluded prior to statistical analysis. Ratios demonstrating a p-value of <0.05 and an adjusted p-value of <0.25 were deemed statistically significant. The statistical significance was evaluated employing the Mann-Whitney U test.

### Consensus Molecular Subtyping

To estimate CMS (Consensus Molecular Subtyping) distribution relative to different treatment outcome machine-learning based algorithms CMSclassifier v1.0.0 ^5^, CMScaller v2.0.1 ^53^ and CMSFFPE v0.1.0 ^54^(specifically developed for FFPE samples) were employed. To ensure the reliability of the results, algorithm outputs were only assessed for samples exhibiting comparable classifications across different algorithms. All methods reported both predicted and nearest classification, while CMSclassifier additionally used two methods, RF (random forest) and SS (single sample) predictor. The difference in distribution of CMS categories among treatment outcomes was evaluated by using the Chi-square/Fisher exact test statistics.

### Gene Set Enrichment Analysis

Gene set enrichment analysis (GSEA) was conducted by using molecular signature database libraries C3 (Gene Ontology Biological Processes), KEGG, Canonical Pathways (Reactome subset) alongside MSigDB databases utilizing packages clusterProfiler v4.10.1^55^ and org.Hs.eg.db v3.18.0 ^56^. Pathways were deemed significant when exhibiting adjusted p-value<0.05, FDR<0.25 and NES>1.5 for those upregulated in pCR and R groups, and NES < - 1.5 for those upregulated in Non pCR and NR groups.

### Analysis of transcription factor activity

Transcription factors (TFs) target genes were retrieved from CollecTRI-derived regulons, composed of data from 12 different sources. An analysis of correlation between treatment response and transcription factor activity was conducted employing the decoupleR package v2.12.0 ^57^ which uses methods based on sign and weight of network interaction. Data input included DESeq2 statistical output, LogFC/se, derived from a combined dataset between R/NR and pCR/Non pCR. To determine the transcription factor activity score a univariate linear model (ULM) was used, while viper and norm_fgsea and norm_wmeans and norm_wsum served as additional validation chose based on Yashar et al results ^58^. Transcriptional factors with statistically significant scores through all methods were considered as significant. In order to determine the involvement of activated transcription factors in signaling pathways, an overrepresentation analysis was performed using the Gene Ontology database of biological processes as a reference. Finally, the correlation of activated TFs and miRNAs, which proved to be potential therapeutic targets in rectal cancer, was investigated using multiMiR v1.24.0 package ^59^.

## RESULTS

We assembled an RNAseq cohort of 248 publicly available transcriptomics data from patients diagnosed with rectal cancer categorized as pathological complete responders (pCR, n = 60) or non-complete pathological responders (Non pCR, n = 188), including 156 samples classified as responders (R, n = 92) and non-responders (NR, n = 64). Patients characteristics are shown in table 4. No differences were observed in the distribution of sex among response groups, as tested with Pearson Chi-square test and pCR/Non pCR groups. The cohort’s age followed normal distribution among R/NR and pCR/Non pCR. A principal component analysis (PCA) plot showing the effectiveness of neutralizing the batch effect associated with the study variable is shown in Figure 3.

**Table 3.**
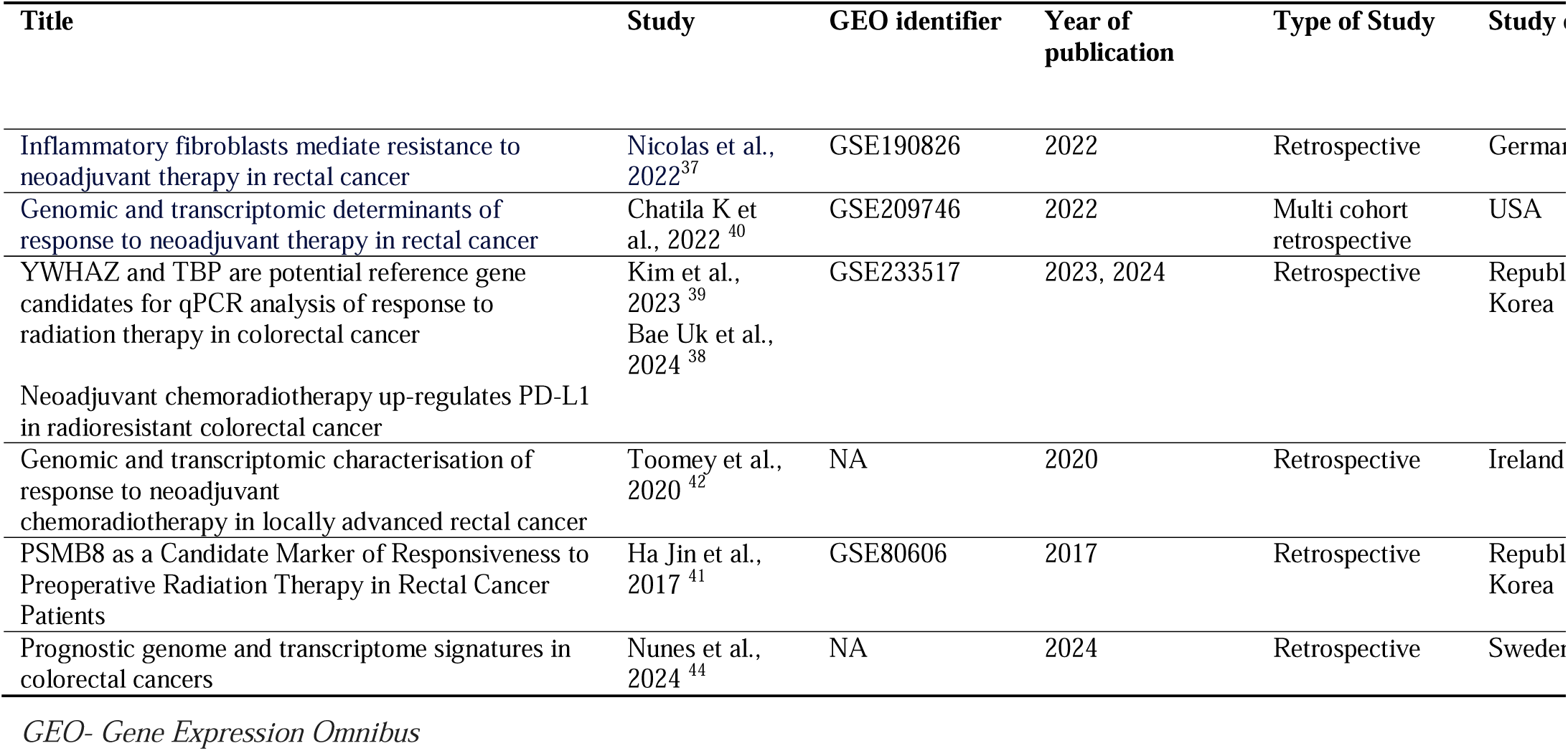
Studies included in meta-analysis.

**Table 4.**
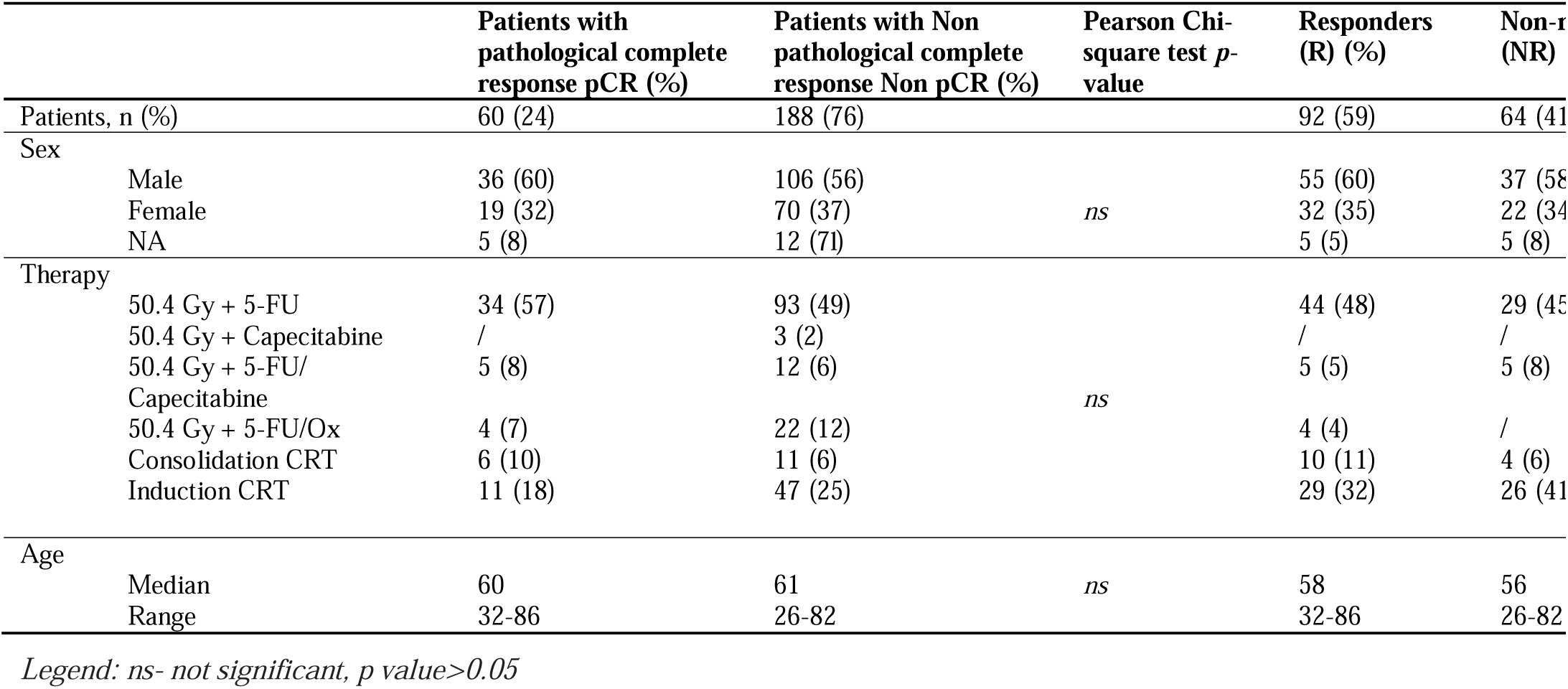
Patient characteristics stratified by therapy response.

### Identification of predictive biomarkers of response to nCRT in LARC

An initial screening for profiling gene expression alterations between response groups was performed both at the individual dataset level, and at the batch-free combined datasets. The combined cohort was composed of four individual datasets containing 17342 and 17510 common genes intersecting between all samples with available R/NR and pCR/Non pCR metadata. Results of differential expression analysis for the individual datasets are shown in Table 5. Figures 4 and 5 demonstrate results from the analysis performed using the combined cohort.

**Table 5.** Results of individual studies and combined dataset.

| Dataset | Number of samples | Number of common genes | p < 0.05 and Log2FC < -1 | p < 0.05 and Log2FC < 0 | p < 0.05 and Log2FC > 1 | p < 0.05 and Log2FC |
| --- | --- | --- | --- | --- | --- | --- |
|  | *R vs NR* |  | *Up in NR* | *Up in NR* | *Up in R* | *Up in R* |
| GSE233517 | 11 (4 R, 7 NR) | 37734 | 330 | 489 | 262 | 357 |
| GSE209746 | 87 (48 R, 39 NR) | 37734 | 219 | 1338 | 172 | 1532 |
| GSE80606 | 22 (9 R, 13 NR) | 37734 | 323 | 490 | 231 | 570 |
| Toomey et al | 10 (5 R, 5 NR) | 33908 | 211 | 240 | 156 | 215 |
| Combined dataset | 156 (92 R, 64 NR) | 17342 | 81 | 700 | 97 | 757 |
|  | *pCR vs Non pCR* |  | *Up in Non pCR* | *Up in Non pCR* | *Up in pCR* | *Up in pCR* |
| GSE233517 | 11 (8 pCR, 3 Non pCR) | 37734 | 233 | 427 | 212 | 276 |
| GSE209746 | 93 (23 pCR, 70 Non pCR) | 37734 | 195 | 1344 | 234 | 2208 |
| GSE80606 | 22 (3 pCR, 19 Non pCR) | 37734 | 2022 | 2599 | 479 | 921 |
| Toomey et al | 17 (5 pCR, 12 Non pCR) | 33908 | 364 | 656 | 343 | 948 |
| GSE190826 | 104 (26 pCR, 78 Non pCR) | 34784 | 330 | 487 | 262 | 674 |
| Combined dataset | 248 (60 pCR, 188 Non pCR) | 17510 | 15 | 329 | 71 | 1316 |

*R*- responder, *NR*- non-responder, *pCR* - pathological complete response, *Non pCR*-Pathological non-complete response, *Log2FC*- *Log2* fold change

**Figure 4.**
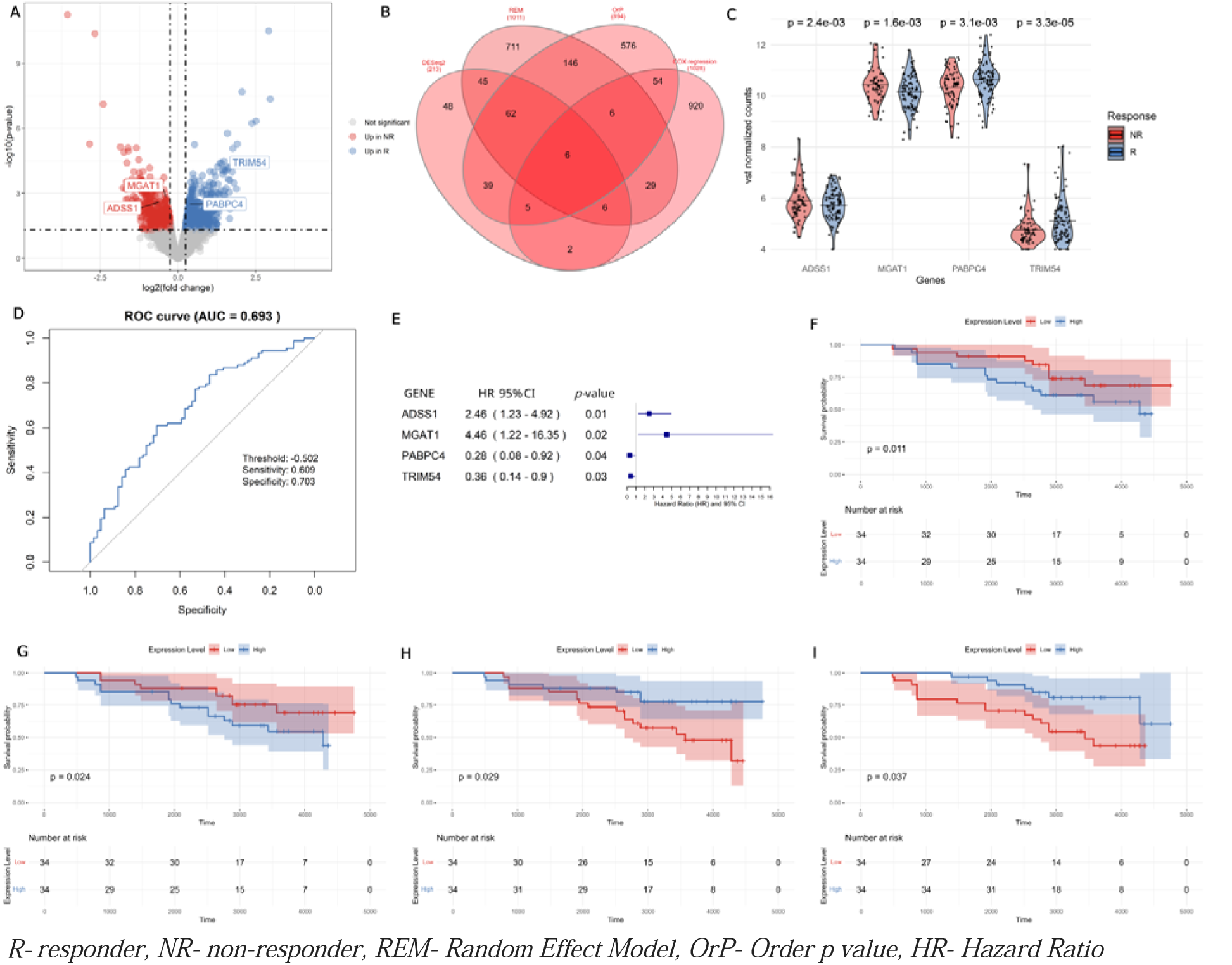
Predictive biomarkers of poor/good treatment response A.Volcano plot of DEGs in R/NR. B. Venn Diagram ^60^ of REM, OrP, DESeq2 and COX regression significant results C.Boxplot of gene expression identified as predictive markers of good response to therapy along with p values from the combined dataset. D. ROC curve obtained from VST normalized gene expression of predictive biomarkers of response to therapy. The combined score was obtained by adding the expression levels of genes overexpressed in good response to therapy, from which the expression level of genes overexpressed in poor therapeutic response was subtracted. E. Forest plot based on univariate COX regression analysis using RFS data and expression level of predictive biomarkers. F/G/H/I. Kaplan-Meier curve of mean expression of *ADSS1*, *MGAT1*, *TRIM54* and *PABPC4* across samples using RFS data. DEGs, differentially expressed genes.

**Figure 5.**
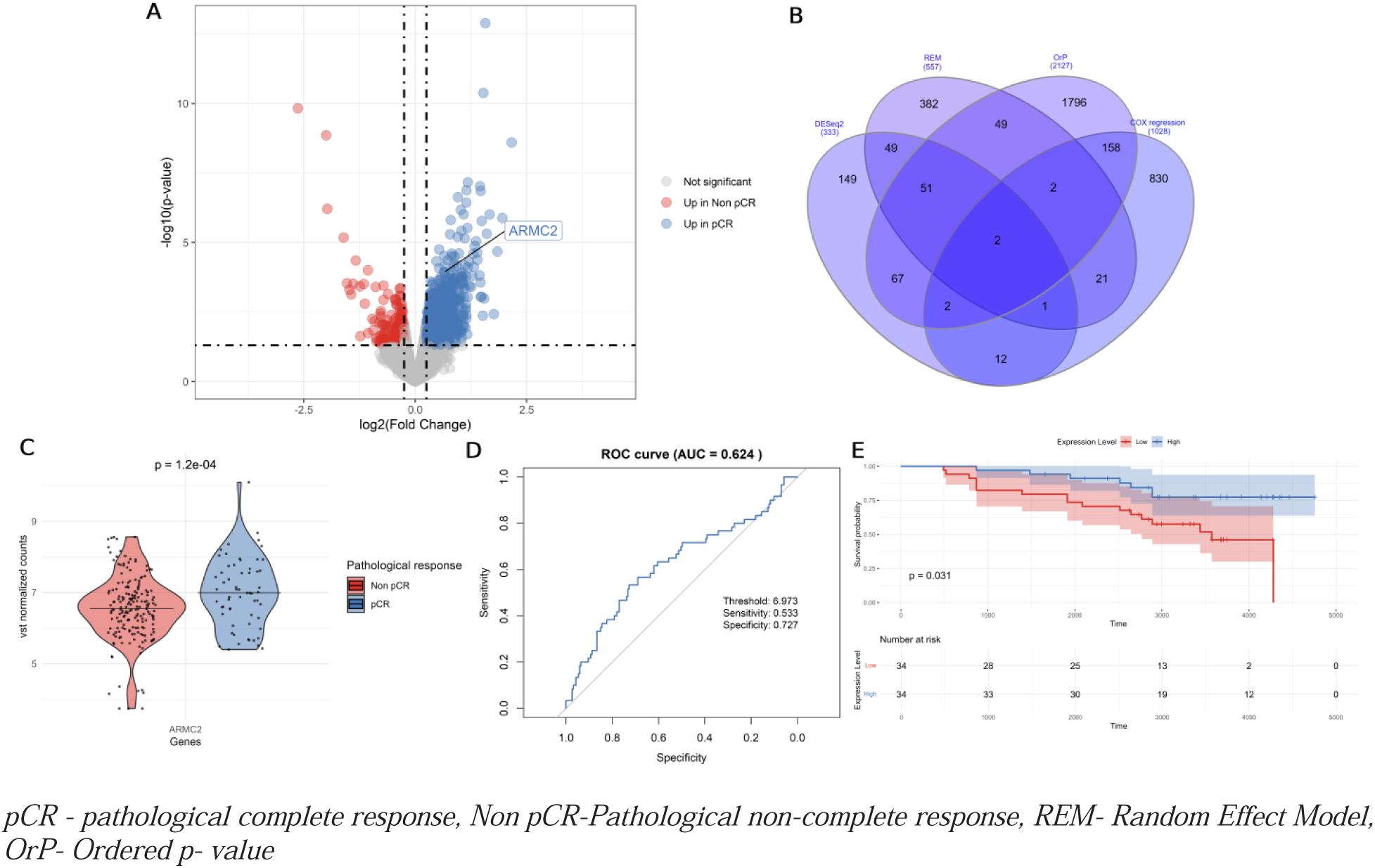
Predictive biomarker of pathological complete treatment response A. Volcano plot of DEGs. B. Venn Diagram ^60^ of REM, OrP, DESeq2 and COX regression significant results C. Boxplot of gene expression identified as predictive markers of pathological complete response to therapy along with p values from the combined dataset. D. ROC curve obtained from VST normalized gene expression of predictive biomarkers of response to therapy. E. Kaplan-Meier curve of mean expression of *ARMC2* across samples using RFS data.

In order to discover predictive markers of response to neoadjuvant chemoradiotherapy we integrated the differential gene expression lists of the combined and the individual datasets using a combination of ordered p-value and a random effect model-based *on* rank aggregation (Supplementary Data 1). Combining studies increased group heterogeneity and statistical power while technical variation was a challenge. On the other hand, by applying meta-analysis, each study is analyzed separately, on the basis of which p-values and effect sizes are estimated for each gene ^61^. By combining these two methods, the advantages of both were used in the identification of predictive biomarkers of response. In order to achieve the maximum number of patients in the discovery cohort, we used a dataset for validation that did not contain information on response to therapy but did have information on time to disease recurrence after surgery. It was expected that patients who had a pathological complete response or a better response would have a longer period of relapse of the disease compared to patients in whom this response was absent. After the analysis of the results, 6 genes were identified that showed significance in the prediction of good and bad therapeutic response. Based on the prognostic potential of the identified genes, it was established that two genes have the potential of a good prognostic factor and are predictive markers of a good response (*TRIM54* and *PABPC4*), while two genes were concluded to be poor prognostic markers and predictors of a poor therapeutic response (*ADSS1* and *MGAT1*). The two markers had opposite predictive and prognostic effects so they were not identified as promising candidates (Table 6, Figure 4). Beyond the predictive biomarkers of R/NR, predictive biomarkers of pathological complete response were investigated. By using the same meta-analysis approach and combining dataset statistics together with RFS data as a source of independent validation two genes were identified as significant but only *ARMC2* were identified as predictive biomarker upregulated in patients with pathological complete response and good prognostic marker in all investigated methods (Table 7, Figure 5).

**Table 6.**
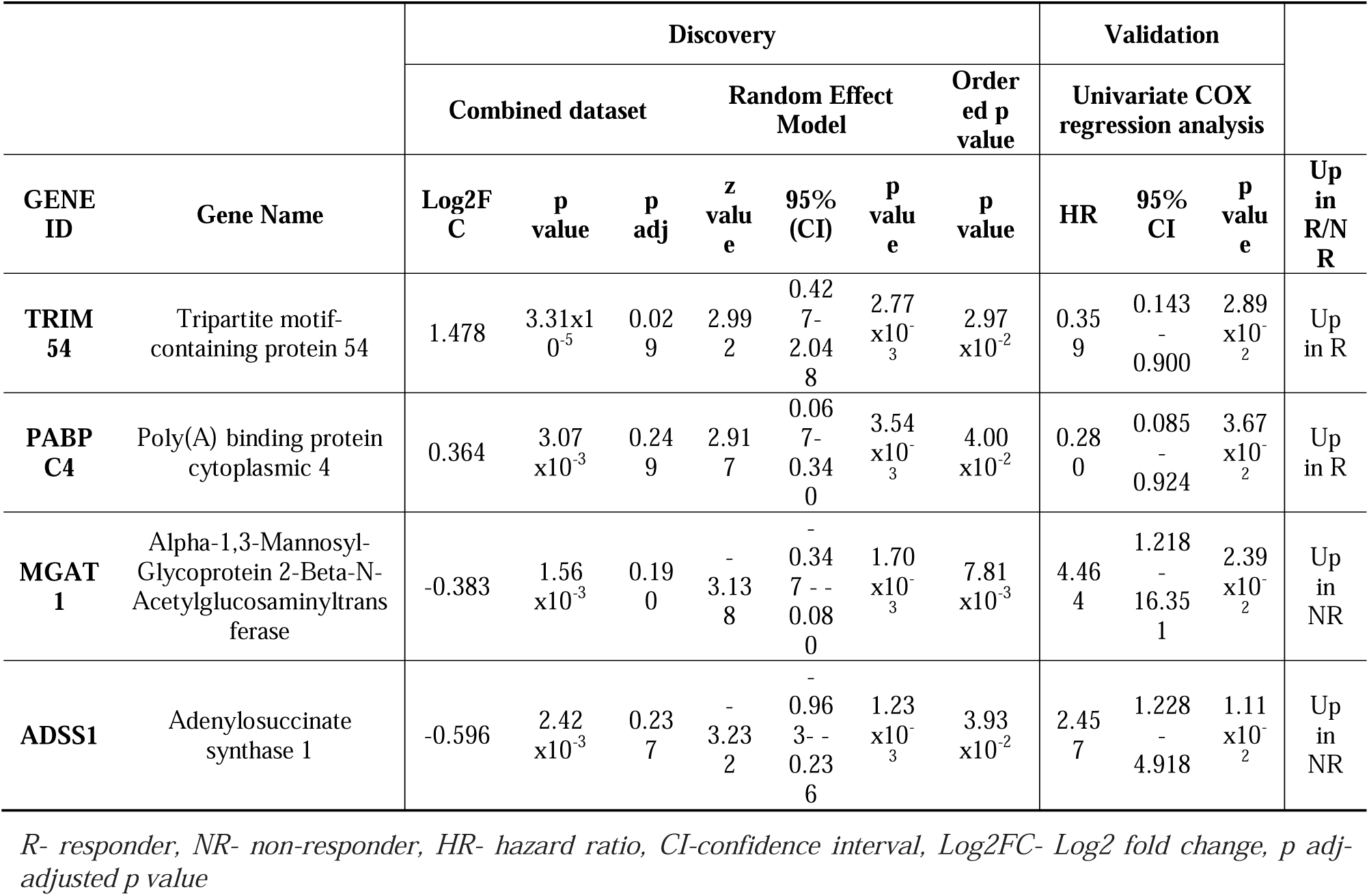
Predictive biomarker of treatment response.

**Table 7.**
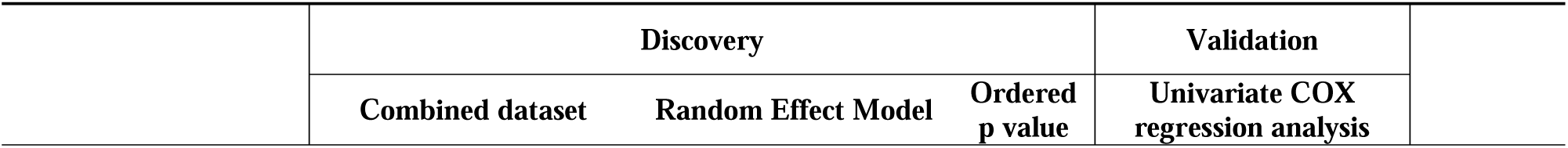
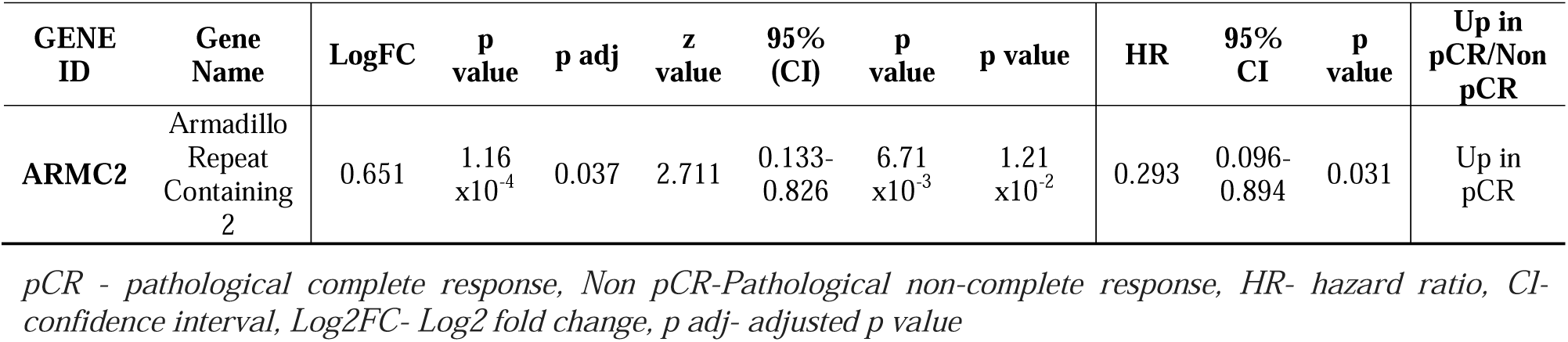
Predictive biomarker of pathological complete treatment response.

Investigation of expression patterns of the top 100 DEGs showed difficulties in predictive capacity concerning heatmap. Covariates such as sex, CMS and microsatellite stability/instability (MSS/MSI) were employed (Figure 6). Further investigation using immune deconvolution methods, gene set enrichment analysis and molecular classification was performed in order to understand underlying mechanisms which effect will be seen further on phenotypic profile.

**Figure 6.**
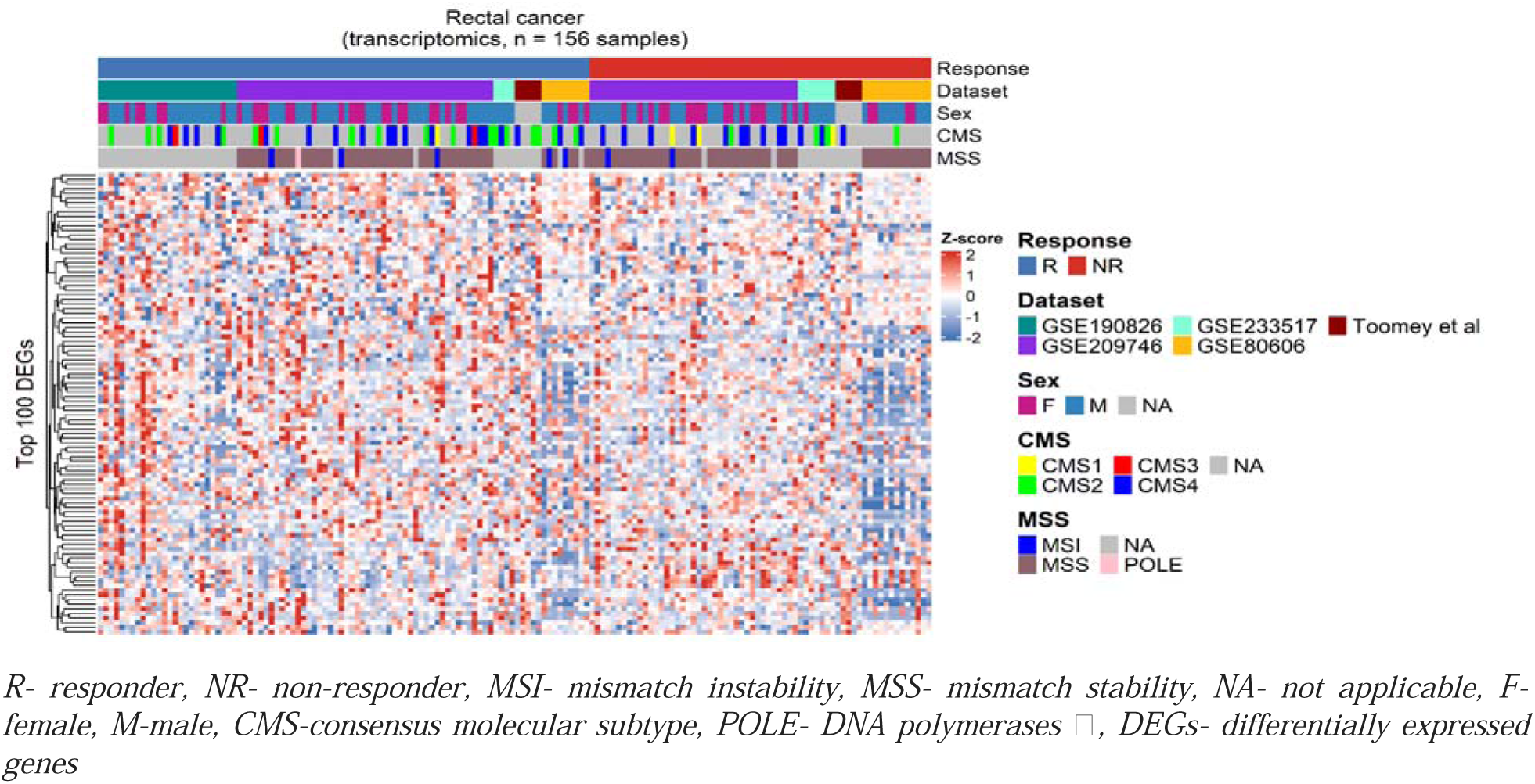
A. Heatmap of Top100 DEGs between R/NR.

### Variability in immune landscape as predictor of therapy response

Immune cell deconvolution from bulk RNA sequencing data was executed utilizing various deconvolution methodologies, including CIBERSORT, xCell, TRIMER, and MCP-counter. The immune deconvolution approaches revealed discrepancies between responders and nonresponders regarding the presentation of immune cells prior to therapy, suggesting activation of the immune system in both protective and suppressive modalities. Notably, the responder cohort exhibited a markedly higher proportion of CD4+ T cells, monocytes, and resting NK cells, thereby enhancing the antitumor environment and facilitating improved treatment outcomes (Table 8). Furthermore, overrepresentation of macrophages, activated myeloid dendritic cells, hematopoietic stem cells, NK T cells, and common lymphoid progenitors were common among patients experiencing poor treatment outcome ^62^. However, employed immune deconvolution methods did not yield identifiable features that would enable prediction of pathological complete response.

**Table 8.** Immune subtypes and immune cell ratio with predictive R/NR capacity.

| Immune cell subtype | Deconvolution Method | p value | p adjusted | Log2 Fold Change | Up in R/NR |
| --- | --- | --- | --- | --- | --- |
| Myeloid dendritic cell activated | xCell | 0.034 | 0.239 | -0.039 | Up in NR |
| T cell CD4+ (non-regulatory) | xCell | 0.030 | 0.239 | 2.84E-04 | Up in R |
| Common lymphoid progenitor | xCell | 0.004 | 0.070 | -0.062 | Up in NR |
| Hematopoietic stem cell | xCell | 0.037 | 0.239 | -0.026 | Up in NR |
| Macrophage | xCell | 0.022 | 0.239 | -0.008 | Up in NR |
| T cell NK | xCell | 0.001 | 0.021 | -0.075 | Up in NR |
| T cell CD4+ | TIMER | 0.006 | 0.037 | 0.039 | Up in R |
| NK cell | MCP counter | 0.001 | 0.009 | 1.555 | Up in R |
| T cell CD4+/Myeloid dendritic cell | TIMER | 0.010 | 0.153 | 0.056 | Up in R |
| T cell CD4+/T cell CD8+ | TIMER | 0.033 | 0.248 | 0.278 | Up in R |
| NK cells resting/Macrophages M1 | CIBERSORT | 0.001 | 0.127 | 2.168 | Up in R |
| NK cell/Endothelial cell | MCP counter | 0.002 | 0.035 | 0.130 | Up in R |
| NK cell/Monocyte | MCP counter | 0.003 | 0.035 | 0.114 | Up in R |
| NK cell/Macrophage/Monocyte | MCP counter | 0.003 | 0.035 | 0.114 | Up in R |
| T cell CD8+/NK cell | MCP counter | 0.003 | 0.035 | -0.057 | Up in NR |
| NK cell/Myeloid dendritic cell | MCP counter | 0.003 | 0.035 | 0.459 | Up in R |
| T cell/NK cell | MCP counter | 0.007 | 0.059 | -2.379 | Up in NR |
| NK cell/Neutrophil | MCP counter | 0.008 | 0.059 | 0.080 | Up in R |
| NK cell/Cancer associated fibroblast | MCP counter | 0.016 | 0.109 | 0.016 | Up in R |
| Cytotoxicity score/NK cell | MCP counter | 0.032 | 0.195 | -1.832 | Up in NR |
| NK cell/B cell | MCP counter | 0.041 | 0.222 | 0.229 | Up in R |

Furthermore, ratios of various immune cells were examined. An increased quantity of NK cells, in comparison to other immune cells, correlates with favorable treatment outcomes, including Endothelial cells, Monocytes, Myeloid dendritic cells, Neutrophils, Cancer-associated fibroblasts, B cells, and resting NK cells/Macrophages. The overrepresentation of CD4+ T cells relative to Myeloid dendritic cells and CD8+ T cells also contributes to improved treatment outcomes. Conversely, poor treatment outcomes were characterized by an overrepresentation of T cells, CD8+ T cells, and cytotoxicity score over NK cells (Table 8, Figure 7).

**Figure 7.**
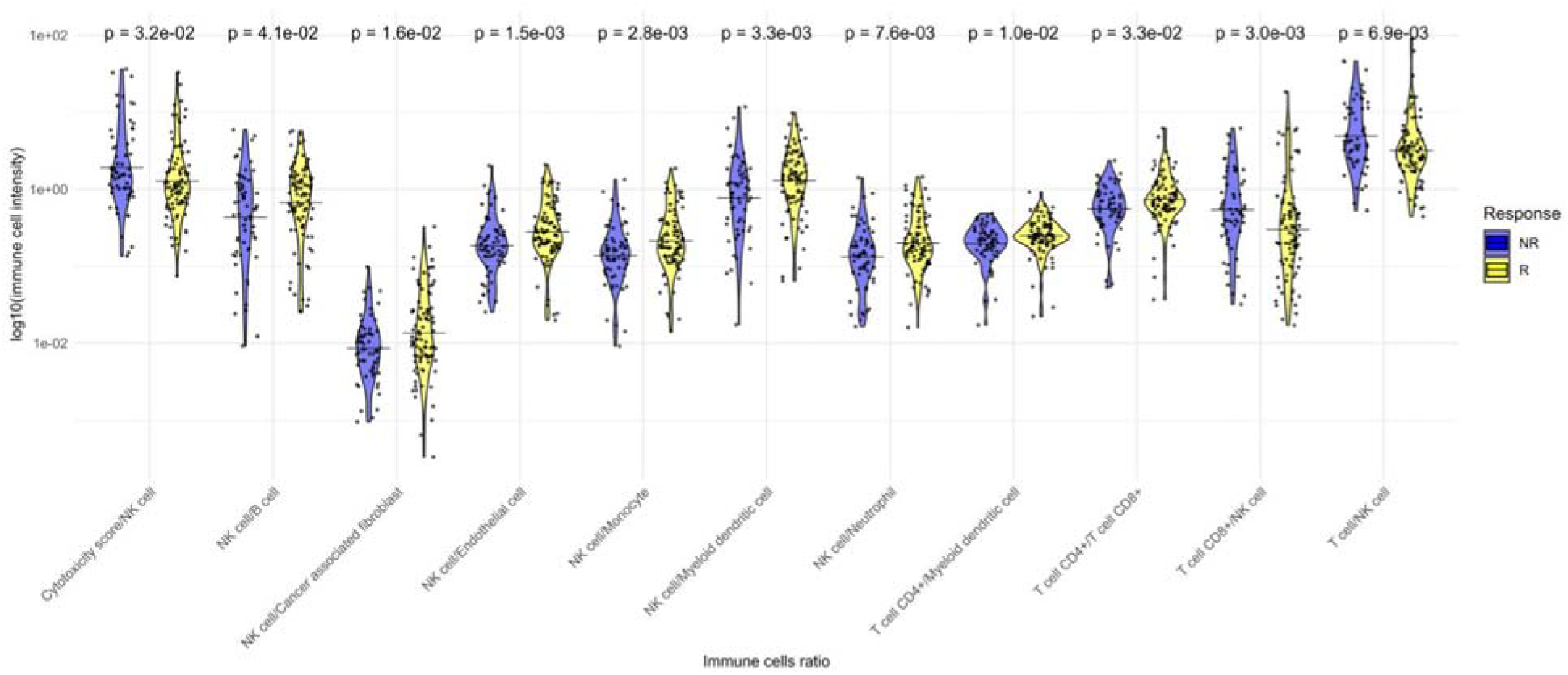
Log 10 Immune cell intensity of significant immune cell ratio between R and NR.

### Molecular subtype-based characterization of tumor response heterogeneity

Different algorithms were used for CMS sample classification. The results were compared and only samples that had consistency in classification between different algorithms were used further for analysis. 51 R/NR and 93 pCR/Non pCR CMS categories were successfully identified (Supplementary Data 2). There is a trend towards a difference in CMS distribution between R and NR (p=0.077). The CMS2 and CMS4 subtypes (Table 7) had higher prevalence R/NR and pCR/Non pCR than the CMS1 and CMS3 subtypes. The PCA plot clearly demonstrates distinct molecular clustering for CMS2 and CMS4 based on transcriptomic profiles (Figure 8D). Patients within the NR category were predominantly classified as CMS4, whereas those in the R category were categorized as either CMS2 or CMS4. Among other CMS categories CMS1 were dominantly characterized as NR, while none of NR were classified as CMS3 (Table 9, Figure 8). Considering pathological complete response most of the patients were identified as CMS2 or 4. Non pCR were dominant over pCR in the CMS3 subtype (Table 9, Figure 8).

**Figure 8.**
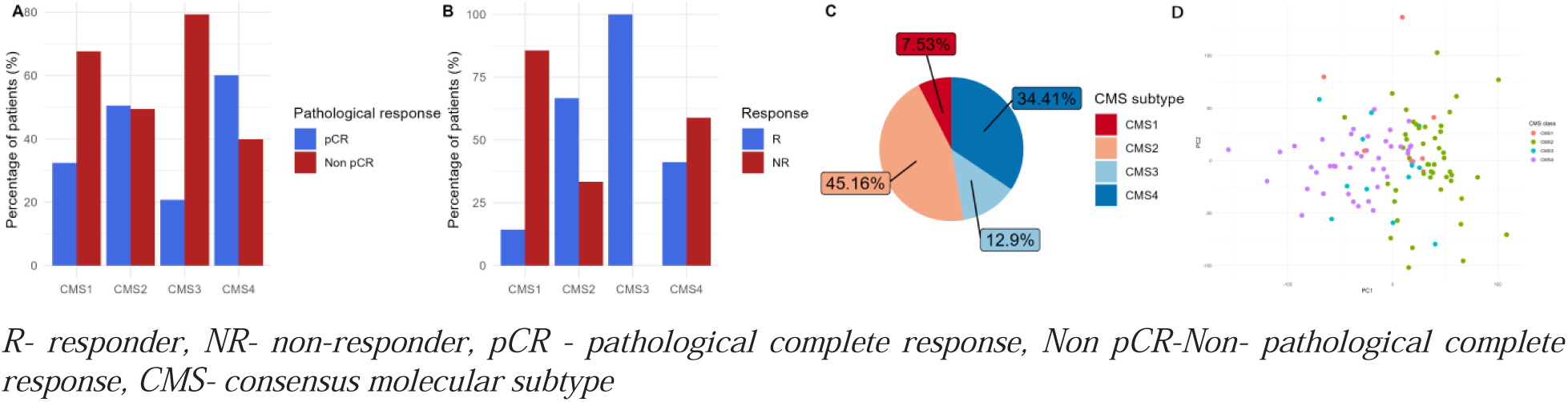
CMS distribution A. and B. Percentage of patients per CMS category scaled by the total number of patients per group (pCR/Non pCR and R/NR). C. CMS subtype distribution of rectal cancer samples. D. PCA plot of transcriptomic data based on different CMS

**Table 9.** Distribution of samples among CMS subtypes.

|  | R | NR | pCR | Non pCR |
| --- | --- | --- | --- | --- |
| **Total** | 34 (100) | 17 (100) | 24 (100) | 69 (100) |
| **CMS1** | 1 (2.94) | 3 (17.65) | 1 (4.17) | 6 (8.70) |
| **CMS2** | 16 (47.06) | 4 (23.53) | 11 (45.83) | 31 (44.93) |
| **CMS3** | 3 (8.82) | 0 (0.00) | 1 (4.17) | 11 (15.94) |
| **CMS4** | 14 (41.18) | 10 (58.82) | 11 (45.83) | 21 (30.43) |

*R- responder, NR- non-responder, pCR - pathological complete response, Non pCR-Pathological non-complete response, CMS- consensus molecular subtype*

### Gene Set Enrichment Analysis highlights differentially activated pathways

Various signaling pathways were enriched in the nonresponder group, primarily related to immune response, necrosis, cell metastasis, movement through extracellular matrix, angiogenesis and vessels development, lipids and lipoprotein metabolism, signal transduction, and many bacterial/viral infection responses (Figure 9). The NR group was characterized by a very complex list of immune-related pathways and Revigo (http://revigo.irb.hr/) ^63^ was utilized to condense these pathways by eliminating redundant GO terms together with Cytoscape ^64^ (Figure 9B). Patients with favourable treatment response showed enrichment in signaling pathways related to cytoskeleton organization, control of transcription, translation, signaling pathways that reduce the occurrence of errors, control of ribosomal RNA processing, translational imprinting, as well as pathways related to piRNA processing, transposons and retrotransposons (Supplementary Data 3, Figure 9). Enriched pathways among patients with pathological complete response showed activation of immune related signaling pathways particularly those related to the activity of B cells and NK cells. The gene set enrichment analysis highlighted the significance of protein synthesis and translation control in patients with Non pCR. This observation is supported by the fact that aside from protein folding control and translation control, enriched signaling pathways include many of those related to energy metabolism. Furthermore, many signaling pathways related to protein synthesis include pathways and processes in endoplasmic reticulum that are usually producing membrane bound proteins as well as proteins that are part of lysosomes, endosomes and secretory vesicles (Supplementary Data 4).

**Figure 9.**
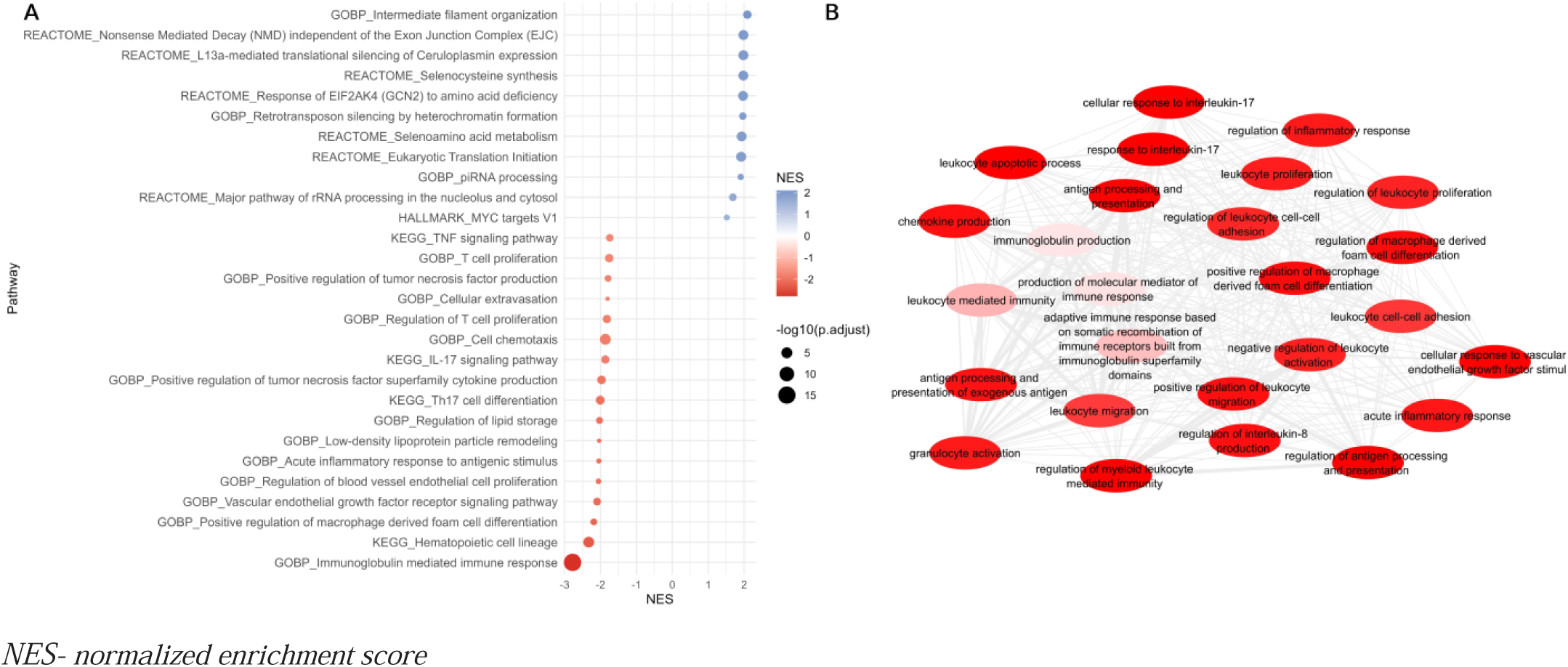
Representative signaling pathways differentially activated between R and NR. A. Selected pathway from Gene Set Enrichment analysis upregulated in R (NES>1.5, blue) and NR (NES< -1.5, red). B. Overview of Immune related pathways enriched in NR group

### Identification of key transcription factors of therapeutic sensitivity and resistance

Since transcription factors can regulate a large number of genes and in that way affect various biological processes, the subsequent phase of the analysis involved a comprehensive examination of transcription factors (TFs). The regulatory functions of TFs exhibit both positive and negative impacts within the responder (R) and non-responder (NR) groups. As illustrated in the volcano plot (Figure 10D,E,F), the number of genes regulated in favor of a particular outcome relative to others dictates their overall significance score. The ULM algorithm was employed to determine the activity of transcription factors based on therapeutic response (Figure 10A), while additional algorithms VIPER, wsum, wmean, and FGSEA were used to validate the obtained findings. A total of 104 TFs were identified as statistically significant (p-value ≤ 0.05) within our studied cohort, which includes 6 enriched in R and 98 in NR. Out of all identified significant TFs, 42 of them were validated by incorporating additional methodologies for evaluating transcription factor activity (wmean, wsum, fgsea, and viper) (Figure 10A/B/C) (Supplementary Data 5). Among NR, the highest statistical significance was found for *SP1* (Figure 11D) and *NFKB* (Figure 10E) while *TCF15* (Figure 10R) was dominant among R.

**Figure 10.**
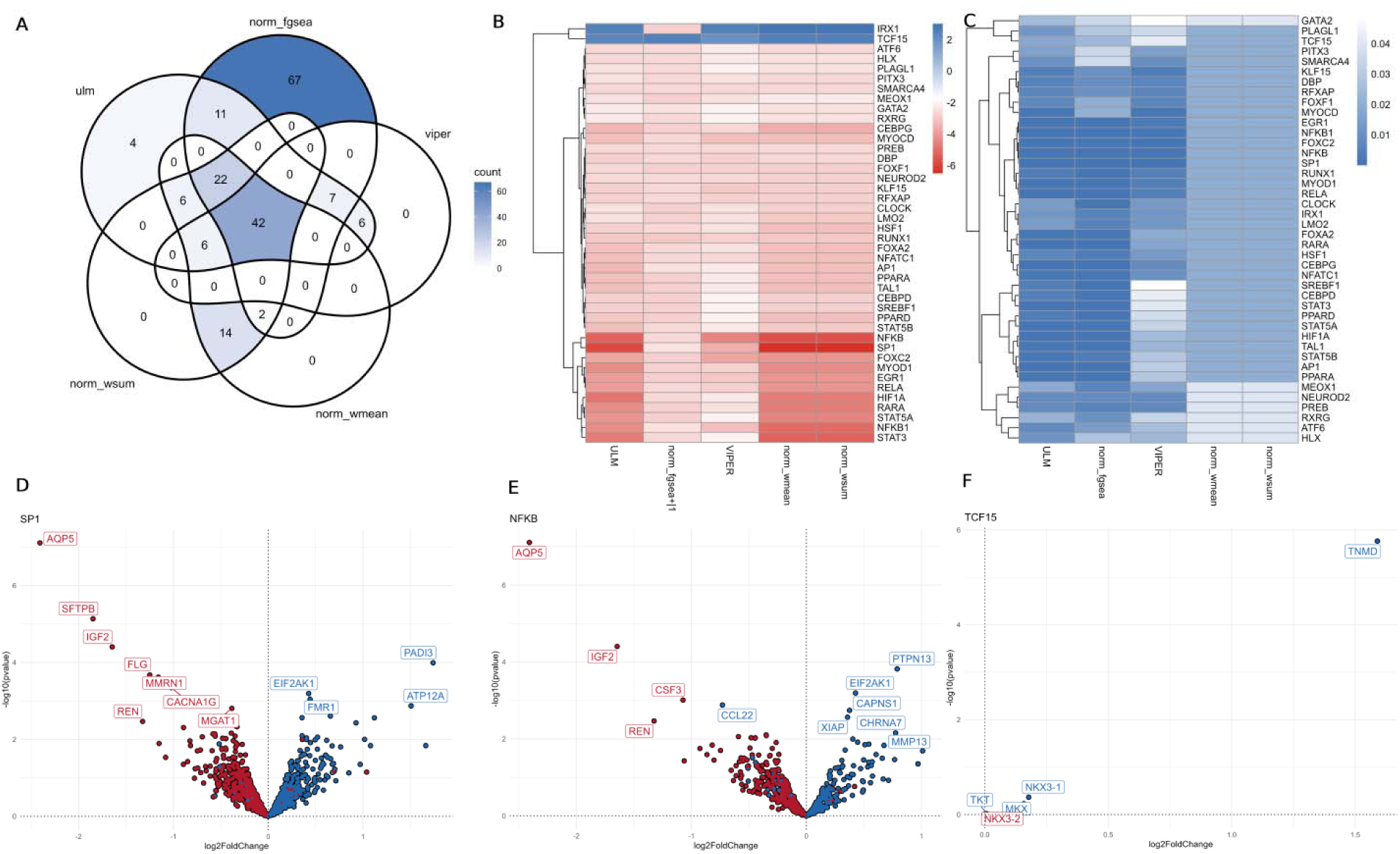
Results of TFs activity among R/NR groups A. Cross section of transcription factors identified by ULM, wmax,wmean, VIPER and FGSEA within R/NR group. B. Score and C. p value of confirmed transcription factors. D/E/F. Volcano plot of DEGs (log2FoldChange vs - log10 P value) with top TFs (D. *SP1*, E. *NFKB* and F. *TCF15*) target genes colored based on TFs effect on them (red is activating effect and blue is suppressive effect)

**Figure 11.**
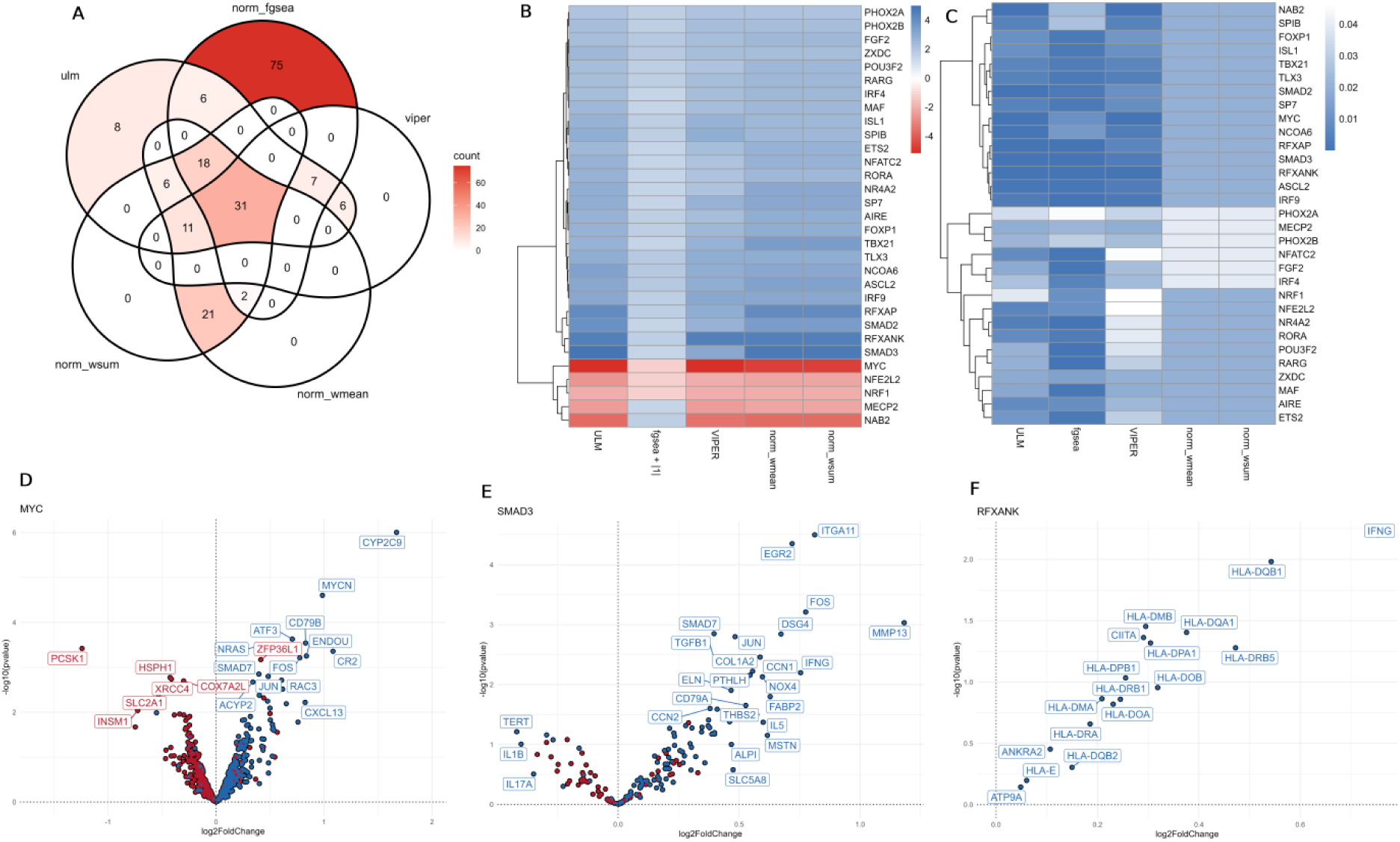
Results of TFs activity among pCR/Non pCR groups A. Cross section of transcription factors identified by ULM, wmax, wmean, VIPER and FGSEA B/C. Score (B) and log10 p-value (C) of overlapped transcription factors. E/F/G. Volcano plot of DEGs (log2FoldChange vs -log10 P value) with top TFs based on score (D. *MYC*, E. *SMAD3*, F. *RFXANK*); target genes colored based on TFs effect on them (red is activating effect and blue is suppressive effect).

An analysis of transcription factors’ activity within the cohort pCR/Non pCR identified 93 TFs with statistically significant activity; of these, 72 TFs had increased activity toward pathological complete response. Through the incorporation of supplementary validation methods, a total of 29 transcription factors were confirmed (Supplementary Data 5, Figure 11A/B/C), 26 of which were activated in favor of a pathological complete response. *SMAD3* and *RFXANK* (Figure 11 F/G) had the most significant effects in favor of pCR. While the most notable effect in favor of Non pCR had *MYC* (Figure 11E). TF upregulated in pCR had a strong specificity toward activating genes upregulated in pCR.

Gene Ontology database was used as a source of signaling pathways and genes involved, used to identify overrepresented pathways among confirmed TFs. Analysis of TFs activated in NR highlights the importance of signaling pathways related to miRNA and ncRNA metabolism and activity (Figure 12A). Furthermore, TFs activated in pCR were part of signaling pathway related to immune response, particularly, CD4+ T cell activation, Th17 immune response and alpha-beta T-cells (Figure 12B).

**Figure 12.**
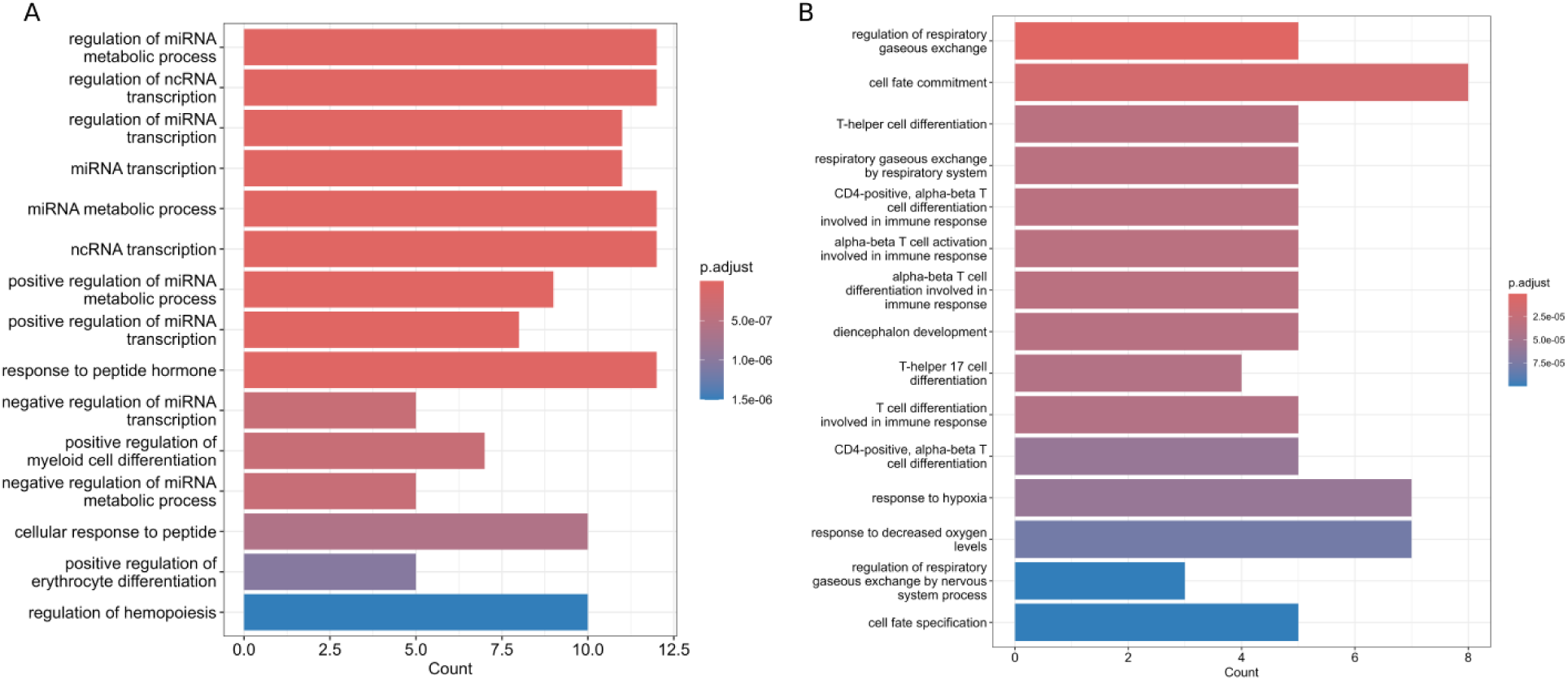
Top 15 signaling pathways overrepresented among TFs differentially activated between R and NR (A) and pCR and Non pCR **(B)**

The review article by Imedio et al. ^65^ provided an overview of miRNAs identified as possible therapeutic targets in rectal cancer. List of miRNAs includes miR-450-5p^66^, miR-577^67^, miR-375-3p^68^, miR-139-5p ^69^ with therapeutic impact on chemotherapy, miR-130a^70^, miR-145^71^, miR-122-5p ^72^ with therapeutic impact on radiotherapy as well as miR-31^73^, miR-129-5p^74^, miR-144 ^75^, miR-381^76^ and miR 19a-3p^77^. A list of potential targets was used to detect interactions with significantly activated transcription factors. Numerous transcription factors that have been demonstrated to be significant in response to therapy interact with the majority of the highlighted miRNAs (Figure 13). The exact direction of regulation is not known, but the results indicate a clear connection between the activation of transcription factors in R/NR and the regulation of miRNAs, whose potential as therapeutic targets has been reported ^65–69,72,74,77^.

**Figure 13.**
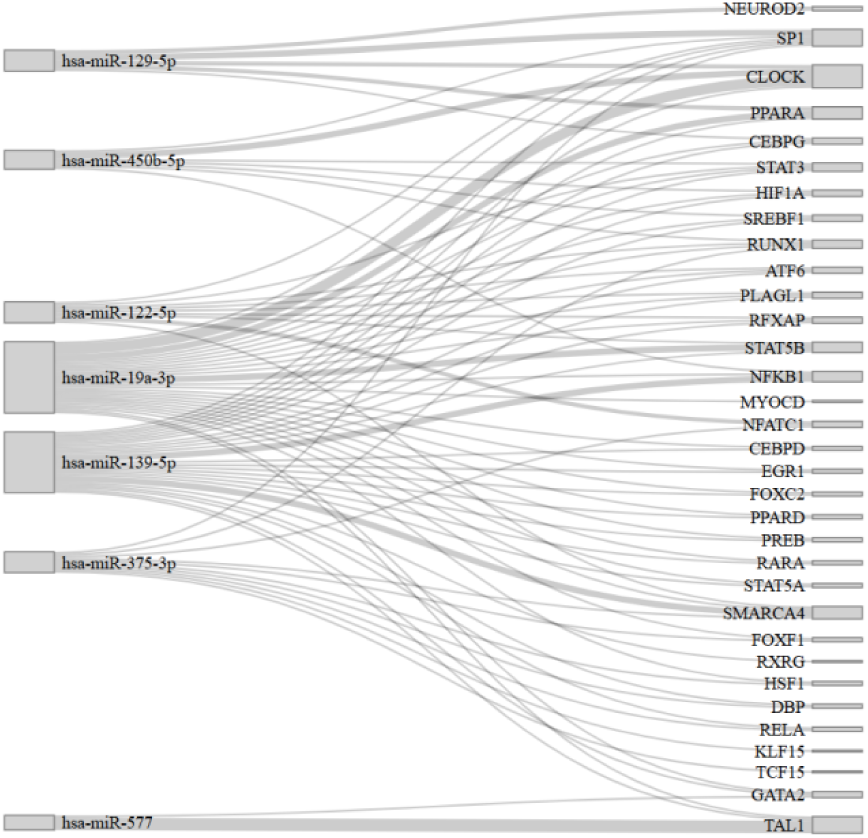
Activated TFs in NR and their targets, miRNAs that have been shown as predictive markers of response to nCRT.

## DISCUSSION

Locally advanced rectal cancer is one of the most commonly diagnosed forms of rectal cancer, with 10–20% of patients achieving a pathological complete response following standard nCRT ^8^. Routine clinical practice and currently available methodologies are insufficient for identifying these patients and providing them with optimized care. Nonetheless, a sizable portion of patients experience suboptimal therapeutic outcomes. This issue is even more critical considering that recent modifications in neoadjuvant treatment — mainly the introduction of additional chemotherapy in the neoadjuvant setting as part of the total neoadjuvant therapy (TNT) approach, and the escalation of radiation doses — are resulting in a greater number of patients achieving a clinical complete response^18–20,22^. Nowadays, in light of these changes in LARC treatment, it is crucial to avoid both overtreatment and undertreatment in order to achieve the best treatment outcomes while preserving patients’ quality of life.

Currently, there are no clinically confirmed predictive markers for response to nCRT, and the precise criteria for identifying patients eligible for the *watch-and-wait* strategy remain unclear. The rationale for this phenomenon is attributed to the often limited patient cohort in studies, alongside the variation in treatment of LARC patients concerning radiation dosage and administration techniques. Furthermore, it is common for available published studies to lack supplementary clinical data and additional omics levels that would facilitate the establishment of a biomarker panel at appropriate levels via a multi-omics approach. Additionally, a significant challenge arises from the population specificity of the identified biomarkers, as their validation across diverse populations is notably absent ^78–80^. Meta-analysis of transcriptional profiles of LARC have provided insight into predictive biomarkers and biological features associated with different treatment outcomes. Combining individual studies revealed higher sample heterogeneity due to different populations and increased statistical power ^61^. It is thus important to synthesize the knowledge obtained from available studies and to proceed to a prospective, multicenter, multinational validation study.

Among the genes examined, two genes were identified with predictive potential of good response to therapy including *TRIM54* and *PABPC4*, up-regulated in responder group, while *ADSS1* and *MGAT1,* up-regulated in non-responder group were identified as predictive biomarkers of poor treatment outcome (Figure 4, Table 5). Notably, these genes have not been previously analyzed as predictive biomarkers for response to neoadjuvant chemoradiotherapy (nCRT) in rectal cancer; however, positive effects of their expression have been documented in other malignancies including colorectal cancer and some of them were recognized as prognostic biomarkers according The Human Protein Atlas (HPA) ^81^.

TRIM54 functions as ubiquitinase associated with FSP1, key ferroptosis regulator, and promotes its ubiquitination-mediated degradation ^82^. *TRIM54* is gene coding inhibitor of *FSP1* thus could promote ferroptosis in rectal cancer and increase cell death rate during radiotherapy resulting in better treatment outcome. The role of TRIM54 as a direct regulator of ferroptosis was described in hepatocellular carcinoma (HCC), where it was shown that Sorafenib modulates FSP1 activity through TRIM54, ultimately inducing ferroptosis ^82^. Although the mechanism of action of TRIM54 is poorly investigated one study on HCC reported that overexpression of TRIM54 lead to Axin1-mediated activation of the Wnt/β-catenin signaling pathway ^83^, which is known, poor prognostic sign in colorectal cancer and is associated with more aggressive tumors ^84^. Although shown to be a poor prognostic factor, *TRIM54* expression in the context of enhancing response to chemotherapy or radiotherapy has not yet been investigated. *PABPC4*, gene coding the poly(A) binding protein cytoplasmic 4, is important RNA-binding protein with role of regulating gene expression. It was not investigated as a predictive biomarker but its role in better survival of CRC patients stage II and stage III was reported ^85^. According to the HPA data in the READ validation dataset, it was shown that LARC patients with higher expression have better survival (*p*=0.036) ^81^. MGAT1 protein coded by *MGAT1* gene, called *mannosyl (alpha-1,3-)-glycoprotein beta-1,2-N-acetylglucosaminyltransferase* is an enzyme involved in the process of N-glycosylation, and according to HPA ^81^, it is a poor prognostic biomarker in stage II and III rectal cancer, which is consistent with the obtained results. *ADSS1*, adenylosuccinate synthase 1, protein product has a role in AMP synthesis thus highly energy-consuming cells like cancer cells can use it to proliferate effectively ^86^. ADSS1 is highly abundant in cardiac and heart tissue, controlling purine de novo biosynthesis but it has low cancer expression level ^81^. According to HPA data, *ADSS1* expression level correlates with long-term survival over 3 years in patients with stage II and III rectal cancer. 5-year survival in the group of patients with high expression is 29% vs. 53% ^81^. Furthermore, predictive biomarkers of pathological complete response genes were investigated, resulting in the identification of *ARMC2*, up-regulated in pCR (Figure 5, Table 6). *ARMC2*, Armadillo repeat containing 2, has a protein product with a role in maintenance of the axonemal central pair complex which is essential for the functionality of cells with cilia^87^. HPA data from both TCGA (*p*=0.026) and Validation cohort (*p*=3.3 × 10) confirmed that *ARMC2* has a prognostic role in rectal cancer stage II/III ^81^. Each of these genes cannot be considered as perfect biomarker but in combination with other monitoring methods including hematological parameters, clinical data, radiomics features or biomarkers on proteomic level, it could be complementary and thus contribute to better stratification of patients for watch-and-wait approach, better local disease control, organ preservation and offer comprehensive disease monitoring.

The examination of phenotypic characteristics in relation to the transcriptional profile is crucial for elucidating the mechanism underlying the response to neoadjuvant chemoradiotherapy (nCRT). Additionally, certain characteristics derived from bioinformatics algorithms may serve as predictive biomarkers of response, subject to validation through molecular genetics methods that would allow their direct measurement ^88–90^. Phenotypic profiling has enabled the identification of immune cell subtypes, the activity of distinct signaling pathways, and the functionality of transcription factors, which vary according to therapeutic response.

Immune deconvolution methods demonstrated that the responder group had a significantly higher proportion of CD4+T cells, monocytes and NK cells which affect the tumor microenvironment and facilitate improved treatment outcomes (Table 8). Existing literature indicates that NK cells are triggered by radiation therapy thus their increased presence in the tumor microenvironment plays a crucial role in improved treatment outcomes and prolonged survival ^91^. Previously conducted *in vitro* and *in vivo* studies highlighted that ionizing radiation increases expression of stress-induced NKG2D ligands like MICA/B and ULBP1–3, making tumor cells more susceptible to NK-cell–mediated killing. Radiotherapy can synergize with NK-based immune responses leading to improved antitumor defense^92,93^. Analyzing the immune cell ratio confirmed that the presence of NK cells in relation to Macrophages M1, endothelial cells, monocytes and macrophages, dendritic cells, B cells, neutrophils and cancer-associated fibroblasts had predictive potential of a good response to therapy in LARC patients. The increased level of CD4+ T lymphocytes relative to myeloid dendritic cells and CD8+ T lymphocytes also had the same effect (Table 8, Figure 7).

Furthermore, available results showed that macrophages, myeloid dendritic cells, hematopoietic stem cells and common lymphoid progenitors are overrepresented within the group of patients with unfavourable treatment outcome (Table 8) ^94^. Most of the cells dominant in patients with a poor therapeutic outcome are precursors in the differentiation of immune cells with specific proinflammatory and tumor suppressor functions. The presence of myeloid progenitor cells and generally immune cell precursors in the circulation and at the tumor site was described as early as the early 2000s, although this is not expected under normal conditions. These cells are thought to have a high potential in suppressing the immune system. It has been shown that these cells can produce anti-inflammatory cytokines including IL-10 and TGFβ and thus reduce the infiltration of T lymphocytes ^95^. In addition, examination of the ratio of immune cells in patients with a poor therapeutic outcome showed an increased presence of CD8+ T lymphocytes compared to natural killer cells (Table 8, Figure 7). As a consequence of the effect of chemoradiotherapy, there may be a process of reducing the expression of MHC class I molecules, which leads to the inability to activate CD8+ T lymphocytes and thus the inability to attack tumor cells, making them ineffective. Together with the decrease in the amount of NK cells, the possibility of anti-tumor defense is absent, which can lead to a poor outcome of the therapy ^96^.

Patients who had favourable response to therapy had enriched signaling pathways related to cytoskeleton organization, transcriptional and translational control, signaling pathways that reduce error occurrence, regulation of ribosomal RNA processing, translational imprinting, alongside pathways related to piRNA processing, transposons and retrotransposons (Figure 9). The enriched signaling pathways imply that treatment-naive tissue has a specific phenotypic profile, resulting in variable responses to therapy among patients. Furthermore, the responder group is predominantly characterized as CMS2 and CMS4, where CMS2 is associated with a proliferative environment and MYC and WNT activation which is in direct correlation with the high transcriptional rate within the cells (Table 9, Figure 8) ^97^. The gene set enrichment analysis identifies signaling pathways enriched within non-responder groups related to immune response, necrosis, cell metastasis and movement through extracellular matrix, angiogenesis and vessels development, lipids and lipoprotein metabolism as well as signal transduction (Figure 9). Collectively, these findings suggest a highly active microenvironment, which correlates with the observation that a majority of patients in the non-responder group are categorized as CMS4, recognized as the most aggressive CMS subtype with enhanced epithelial-mesenchymal transition (EMT) activation, angiogenesis, and matrix remodeling (Table 9, Figure 8) ^97,98^.

Enriched pathways among pCR groups showed activation of immune related signaling pathways especially those related to activity of B cells and NK cells. Patients with pCR are mainly distributed among two molecular categories CMS4 and CMS2. Gene set enrichment analysis highlighted the importance of protein synthesis and translation control in patients with Non pCR. Activation of signaling pathways related to protein synthesis indicates that the key in difference in treatment response could be on the protein level. Controlling translation and protein degradation, cancer cells can promote oncogenic phenotype through selective translation of mRNA transcripts that can be crucial for tumorigenesis, therapy resistance and metastasis, or degrading transcripts of mRNA that protein product would promote cell death ^99^. This is supported by the fact that apart from protein folding control, translation control, enriched signalling pathways include many of them related to energy metabolism. Bearing in mind that protein-synthesis is the most energy consuming process, which is supported by our results, showing that rectal cancer cells in patients with non pathological response have a high rate of energy production that is needed for translation control. In addition, our data support that many alterations in protein synthesis are mainly related to proteins processed at endoplasmic reticulum, proteins that are usually membrane bound, as well as part of lysosomes, endosomes and secretory vesicles. Those proteins could act as mediators to the tumor microenvironment supporting the protective role of cancer cells, but also could be used as a good secretory biomarker or drug target. Molecular classification of patients with Non pCR indicated that most of them are classified as CMS2 and then CMS4, groups that are classified with high proliferative rate and EMT and angiogenesis, followed by CMS3, with highest prevalence of *KRAS* mutations and metabolic deregulation (Table 9, Figure 8) ^97^. EMT has been recognized as a key mechanism contributing to resistance to chemoradiotherapy (CRT) in various cancers, including rectal cancer ^100,101^.

Transcription factor activity analysis identified 42 transcription factors exhibiting alterations in responders/non-responders, confirmed by various models. Prominent transcription factors highlighted the importance of immune response modulation, inflammation in patients with poor therapeutic response (Figure 12). Furthermore, the identified transcription factors are involved in the processes of proliferation, differentiation, cellular response to stress, hypoxia, DNA damage, oncogenesis and tumor growth. Among these, *NF-KB* and *SP1* demonstrated the highest score enriched in the NR group and *RCF15* in the R group (Figure 10). It is shown that TFs *NFKB* and *STAT3* are key factors that enable cancer cells to avoid apoptosis and regulate angiogenesis and invasiveness ^102^. The activation of these two transcription factors is critical in linking immunity and tumorigenesis, inducing the expression of proteins associated with angiogenesis, hypoxia, chemokines, and immunosuppressive cytokines ^102^. NF-KB promotes anti-apoptotic signaling (e.g., upregulation of Bcl-2, Survivin), promotes radioresistance through the STING–IFNAR1 axis in colorectal cancer, promotes immune evasion and tumor-promoting inflammation ^103,104^. In research analyzing the effects of FoxO3, a transcription factor regulated by SP1, findings indicated that SP1 enhances FoxO expression which contributes to CRC progression, while knockdown of FoxO3 resulting in lower activity of SP1 lead to reduced cell migration and proliferation ability ^105^. Studies have shown that miR-375-3p can target SP1 and thereby increase the sensitivity of CRC cells to 5-FU-based chemotherapy ^106^. Overrepresentation analysis highlighted involvement of activated TFs in signaling pathways related to miRNA and ncRNA activation and metabolism. Literature data highlight that miRNAs can be used as therapeutic targets. Our results indicate that miR-129-5p, miR-450b-5p, miR-122-5p, miR-19a-3p, miR-139-5p, miR-375-3p as potential therapeutic targets are regulated by transcription factors activated in patients with different therapeutic responses (Figure 13) ^65,107–111^.

The analysis of transcription factors activity within the pCR/Non pCR cohort identified 29 transcription factors mostly enriched within the pCR group. *SMAD3* and *RFXANK* had the most significant effects in favor of pCR. While the most notable effect in favor of Non pCR had *MYC* (Figure 11). *RDXANK* is a key regulator of MHC II complex expression that is required for antigen presentation and activation of CD4+ T lymphocytes ^112^. Although the representation of immune subtypes in patients with complete regression was not significant when it comes to CD4+ T lymphocytes, they were noted to be differentially activated in patients with a better therapeutic outcome. This confirms the positive effect of direct or indirect activity of CD4+ T lymphocytes in response to therapy. *MYC* is a known oncogene and regulator of cell proliferation as well as a key regulator of cell growth while increased activation of *SMAD3* by TGFβ allows inhibition of *MYC* transcription leading to inhibition of proliferation, differentiation and cell death ^113^. Confirmed response modulators are involved in immune response, cell differentiation according to overrepresentation analysis (Figure 12). The modulation of transcription factor activity presents a valuable avenue for in vitro research with potential translational implications. Transcription factors regulate numerous target genes, rendering expression mimicry challenging; however, enhancing expression in patients with phenotypes predisposed to poor therapeutic responses may significantly improve treatment outcomes, thereby contributing substantially to therapeutic efficacy.

Our research group has previously investigated various levels of omics methodologies with the objective of predicting response to neoadjuvant chemoradiotherapy in LARC. Proteomic analysis using DIA-MS/MS ^33^, initial level of hematological parameters ^34^ and a machine learning model based on radiomics of pretreatment magnetic resonance imaging (MRI) 3D T2W contrast sequence scans in conjunction with clinical parameters ^35^ revealed moderate to high predictive potential in term of response to nCRT in LARC patients. The results shown in our studies as well as the results shown in other studies at other molecular and clinical levels indicate that one approach is probably not enough to effectively predict the response to therapy or pre select patients for a watch-and-wait approach. Further studies are necessary to include well-defined cohorts with detailed clinical data and well-molecularly profiled samples (genomic, transcriptomic, proteomic level) with radiomics data, hematological and pathological characteristics that would be used together through machine learning methods to construct a predictive model. In this way, each of the mentioned factors that has the potential of prediction would be additionally supported by other predictive parameters.

Although meta-analysis methods can elucidate significant biological factors that influence different phenotypes, they have limitations. The neutralization of the batch effect in order to reduce the technical variability or unavailability of raw counts undoubtedly affects the biological variability, which is not fully observed by any existing method. Therefore, the results obtained through meta-analyses should be further validated by *in vitro, in vivo and ex vivo* experiments. Additionally, FFPE or tissue biopsy represents only a small portion of the tumor microenvironment, even though it is a gold standard in tumor diagnosis. Furthermore, as the tumor cells from LARC can invade the muscularis propria, which also secretes biological molecules and is not accessible during routine diagnostic analysis, we need to approach other body fluids as complementary sources of information.

## CONCLUSION

The meta-analysis enabled the profiling of the response to neoadjuvant chemoradiotherapy in a geographically heterogeneous but clinically uniform group of patients. It provided insight into the characteristics that enable a better understanding and prediction of the response to therapy. By applying bioinformatics methods and integrating different algorithms, the transcriptional profile of patients with LARC was characterized in detail. In this study, important characteristics of tumors in patients with a poor therapeutic outcome were identified, which can be further investigated through functional studies to design target therapies that would improve the treatment of patients. Markers of a good therapeutic response were also identified, which represent an important contribution to designing a panel of characteristics that would be used for highly effective selection of patients for a wait-and-watch approach to treatment. As the first study of this type, we believe it represents a significant contribution to the current state of the art and a basis for further research, emphasizing the importance of integrating clinical data with molecular data in order to effectively subgroup patients for tailoring treatment depending on expected outcome.

## Supporting information

Supp. Mat. 1

Supp. Mat. 2

Supp. Mat. 3

Supp. Mat. 4

Supp. Mat. 5

## Acknowledgements

This study was funded by the Horizon Europe STEPUPIORS Project (HORIZON-WIDERA-2021-ACCESS-03, European Commission, Agreement No. 101079217). The study was supported by the COST action TRANSLACORE (Translational control in Cancer European Network, European Cooperation in Science and Technology, Action No. CA21154) and the Ministry of Science, Technological Development and Innovation of the Republic of Serbia (Agreement No. 451-03-136/2025-03/200043).

## Author Contributions

A.S. Conceptualization, Data Curation, Formal Analysis, Investigation, Methodology, Visualization, Writing-original draft, Writing-review and editing; R.S. Conceptualization, Data Curation, Formal Analysis, Investigation, Methodology, Writing-review and editing, Supervision; M.M. Conceptualization, Data Curation, Investigation, Methodology, Writing-original draft, Writing-review and editing, Supervision; A.D. Funding acquisition, Investigation, Project Administration, Writing-review and editing; S.S-R. Supervision, Writing-review and editing; R.J. Funding acquisition, Project Administration, Supervision, Writing-review and editing; S.C.B Funding acquisition, Project Administration, Supervision, Writing-review and editing; R.F. Funding acquisition, Project Administration, Supervision, Writing-review and editing; A.V. Funding acquisition, Investigation, Methodology, Project Administration, Supervision, Writing-review and editing; J.Z. Conceptualization, Funding acquisition, Investigation, Methodology, Project Administration, Supervision, Writing-review and editing; M.C Conceptualization, Funding acquisition, Investigation, Methodology, Project Administration, Supervision, Writing-original draft, Writing-review and editing. All authors have read and agreed to the published version of the manuscript.

## Data availability statement

The datasets analyzed in this study are publicly available in the Gene Expression Omnibus (GEO) repository under the accession numbers GSE190826, GSE209746, GSE80606, and GSE233517 (https://www.ncbi.nlm.nih.gov/geo/). An additional dataset used in this study was obtained directly from the corresponding author of the study by Toomey et al. (DOI: 10.3390/cancers12071808) and is available from the authors upon reasonable request.

